# RSV healthcare burden in adults before and since the emergence of the COVID-19 pandemic in 6 European countries

**DOI:** 10.1101/2024.09.20.24314093

**Authors:** Arantxa Urchueguía-Fornes, Richard Osei-Yeboah, Ombeline Jollivet, Caroline Klint Johannesen, Toni Lehtonen, Michiel van Boven, David Gideonse, Rachel A. Cohen, Alejandro Orrico-Sánchez, Rolf Kramer, Thea K. Fischer, Terho Heikkinen, Harish Nair, Harry Campbell, PROMISE investigators

**Affiliations:** Vaccine Research Department, Foundation for the Promotion of Health and Biomedical Research in the Valencian Region (FISABIO), Valencia, Spain; CIBER de Epidemiología y Salud Pública, Instituto de Salud Carlos III; Centre for Global Health, Usher Institute, The University of Edinburgh, Edinburgh, United Kingdom; Sanofi, Lyon, France; Department of Virus and Microbiological Special Diagnostics, Statens Serum Institut, Copenhagen, Denmark; Department of Clinical Research, North Zealand University Hospital, Capital Region, Denmark; Department of Health Security, Finnish Institute for Health and Welfare, Helsinki, Finland; Centre for Infectious Disease Control, National Institute for Public Health and the Environment, the Netherlands; Julius Center for Health Sciences and Primary Care, University Medical Center Utrecht, Utrecht University, Utrecht, the Netherlands; GSK, Rockville, Maryland, United States; Catholic University of Valencia, Valencia, Spain; Department of Pediatrics, University of Turku and Turku University Hospital, Turku, Finland; (University of Edinburgh); (THL); (RIVM); (University Medical Center Utrecht); (Imperial College London); (University of Oxford); (Novavax); (Pfizer); (GlaxoSmithKline); (Janssen); (FISABIO-Public Health); (Sanofi Pasteur); (Teamit)

**Author notes:** **Corresponding author** Richard Osei-Yeboah, PhD, Centre for Global Health, Usher Institute, The University of Edinburgh, 5-7 Little France Drive, BioQuarter Gate 3 - Edinburgh, EH16 4UX, United Kingdom., **Alternate Corresponding author**, Arantxa Urchueguía-Fornes, PhD, Vaccine Research Department, Foundation for the Promotion of Health and Biomedical Research in the Valencian Region (FISABIO), Valencia, Spain. Shared first authorship (Arantxa Urchueguía-Fornes, Richard Osei-Yeboah).

**Keywords:** Respiratory syncytial virus, burden, adults, hospitalisation, surveillance, mortality, ICU admissions

## Abstract

**Introduction:** Respiratory Syncytial Virus (RSV) is a major cause of morbidity in older adults. With the emergence of the coronavirus disease 2019 (COVID-19) and the subsequent changes in respiratory viral circulation, it is crucial to reassess RSV-associated healthcare burden in adults. This study assessed RSV-associated healthcare burden in adults in six European countries before and during the COVID-19 pandemic.

**Methods:** We conducted a retrospective analysis using national hospital admissions data from Denmark, England, Finland, the Netherlands, Scotland, and regional surveillance data from the Valencia region (Spain). We included patients aged ≥18 years hospitalised for respiratory tract infections (RTIs) from 2016 to 2023. We assessed RSV-coded and laboratory-confirmed hospitalisations, intensive care unit (ICU) admissions, in-hospital length of stay (LOS), and mortality.

**Results:** RSV-associated hospitalisations significantly reduced during the 2020/2021 season across all countries, coinciding with strict COVID-19 preventive measures, but resurged in subsequent seasons. We observed the highest hospitalisation rates in adults aged ≥85 years. RSV-coded hospitalisations were found to underestimate the true burden when compared with laboratory-confirmed cases. Underestimation factors ranged from 1.1 to 4.3 times across countries. No significant differences were observed in LOS or ICU admission rates for RSV-associated hospitalisations compared to RTIs.

**Discussion:** Our findings underscore the complex epidemiology of RSV in older adults. The differences between RSV-coded and laboratory-confirmed cases highlight the critical need for improved surveillance and diagnostic practices to better assess the true burden. Our findings could be vital for guiding public health strategies, particularly with the recent introduction of RSV vaccines for older adults.

## Introduction

The impact of respiratory syncytial virus (RSV) in older adults is less understood than in young children. Recent findings show higher hospitalisation rates than previously acknowledged, particularly among adults with underlying health conditions, immunocompromised or of advanced age [1-8]. RSV can exacerbate conditions such as asthma, chronic obstructive pulmonary disease (COPD) or heart failure, leading to severe outcomes, such as pneumonia, myocardial infarction or death [9, 10]. Still, accurately determining the true incidence of RSV-related hospitalisations in older adults is challenging due to a lack of routine RSV testing and dedicated surveillance systems and case definitions [11-14].

With the recent approvals of RSV vaccines for adults ≥60 years of age (Arexvy – GSK, mRESVIA-Moderna and Abrysvo^™^ - Pfizer) [15-17], accurate RSV burden estimates to assist in immunisation strategies and monitor the vaccines’ impact and effectiveness have become a public health priority. The coronavirus disease 2019 (COVID-19) pandemic and associated non-pharmaceutical interventions (NPIs) had significant short-term impacts in lowering some respiratory viral circulations [18, 19]. This reduction is known to have had impact RSV epidemiology including healthcare burden [20, 21]. RSV returned in 2021 and 2022 after COVID-19 measures were lifted, but the patterns of RSV return varied across geographies (e.g., out-of-season peaks, year-round hospitalisations).

This study provides direct estimates of RSV healthcare burden in adults ≥18 years, by estimating changes in frequency and burden of RSV-associated illnesses before and since the COVID-19 pandemic (from 2016 to 2023) in six European countries: Denmark, England, Finland, the Netherlands, Scotland, and the Spain-Valencia region, as part of the PROMISE (Preparing for RSV Immunisation and Surveillance in Europe) initiative [22]. We present findings on RSV-coded and laboratory-confirmed RSV hospitalisation rates, in-hospital length of stay (LOS), RSV intensive care unit (ICU) admissions, and RSV mortality data. We compare admission rates across countries and age groups and provide estimates before and during the COVID-19 era.

## Methods

### Study design

A retrospective study of all-cause respiratory tract infection (RTI) admissions, RSV-coded admissions and RSV laboratory-confirmed admissions using routinely collected hospital admissions data from five countries (Denmark, England, Finland, the Netherlands, and Scotland) and a hospital-based active surveillance network in one country (Spain-Valencia region).

### Study population

National hospital registries containing individual-level patient data on hospital admissions of patients ≥18 years were used for Denmark, England, Finland, the Netherlands, and Scotland. In Spain-Valencia, data from a regional prospective hospital-based active surveillance network was used [23, 24] (see Supplementary Methods). Hospital admissions of patients ≥18 years with any mention of RTI using ICD-9-CM or ICD-10 codes (full list available in Supplementary Material) were extracted in Denmark, England, Finland, the Netherlands, and Scotland. In Spain-Valencia, all hospital admissions for individuals aged ≥18 years, arriving through the emergency room, meeting preliminary inclusion criteria (i.e., residents of the catchment area, not institutionalised and not hospitalised in the previous 30 days) and complying with the influenza-like illness (ILI) [25]) case definition were included (see Supplementary Methods).

### Study period

The study period covered full calendar years from 2016/17 to 2022/23; country-specific available study periods are described in the Supplementary Methods. A year was defined starting from ISO week 27 of one given year to ISO week 26 of the following year. In Spain-Valencia, data were adjusted to the RSV circulation period to allow comparisons across years with different lengths of the active monitoring period (Supplementary Methods). A calendar year is referred to from here on as season.

### Outcomes

Hospital admissions were included if lasting more than 12 hours (8 hours in Spain-Valencia, any duration for England). Scheduled or routine admissions, as well as re-admissions within 28 days (30 days in Spain-Valencia) for the same diagnosis, were excluded. The following hospital admission types were defined: 1) RTI-coded admissions, identified by any ICD-9-CM or ICD-10 code related to a respiratory tract infection (full list of codes in Supplementary Methods) 2) RSV-coded admissions defined as any RTI admission including an RSV-associated ICD-9-CM or ICD-10 code (4801, 46611, 0796, J12.1, J20.5, J21.0, B97.4). 3) RSV-confirmed admissions, where a positive RSV polymerase chain reaction (PCR) test was performed within –7 to +2 days of the hospitalisation (data available only in Denmark, Finland, Scotland, and Spain-Valencia.

A hospital admission was considered completed upon hospital discharge or death. In England, data on ICU admissions were also available. Mortality data (available in England, Finland and Spain-Valencia) included any death during or after hospitalisation where the primary or secondary causes of death included one of the diagnoses described in the Supplementary Methods. For deaths occurring post-discharge (Finland only), those that occurred 14 days after the end of the hospitalisation were excluded. Hospitalisation types were further classified by the following diagnosis groups: upper respiratory tract infection (URTI), lower respiratory tract infection (LRTI) stratified by bronchitis & bronchiolitis, pneumonia & influenza, severe acute respiratory syndrome coronavirus 2 (SARS-CoV-2) or unspecified LRTI (Supplementary Methods).

### Data analysis

Data from patients aged ≥18 years were analysed and stratified by the following age groups: 18-64, 18-49 (only in Finland and Scotland, in Finland denominators were not available for this age group), 50-64 (only in Finland, Scotland, and Spain-Valencia, in Finland no population data available for this age group), 65-74, 75-84 and ≥85 years. Age was calculated as the person’s age at the time of each hospitalisation. Due to data privacy compliance, Denmark, Finland, the Netherlands, and Scotland provided approximate counts when numbers were between 1-4, setting them to 2. Admission numbers were calculated by hospitalisation type, age group, diagnosis group, country, and season, and further stratified by COVID-19 period. We defined before COVID-19 admissions as the average (±SD) of all admissions between 2016/17-2018/19 (2017/18-2018/19 for England and Scotland) and during COVID-19 as the average (±SD) of all admissions between 2021/22-2022/23 (2021/22 only for Denmark and Scotland). For the COVID-19 stratification, we excluded admissions from seasons 2019/20-2020/21, as they were not considered representative seasons due to the strong NPI measures in place. During season 2019/20, 37,462 RTI admissions were coded as SARS-COV-2 in England, 1,351 in Finland, 11,105 in the Netherlands and 3,791 in Scotland (data not shown), hence this season was excluded from the before COVID-19 group in all countries.

ICU admission and in-hospital case fatality rates were calculated using the corresponding hospitalisation numbers as denominators. The in-hospital LOS in days was estimated from admission and discharge dates. It included the ICU stay or the transfer to another hospital, if any. LOS median and interquartile ranges were calculated by hospitalisation type, age group, diagnosis, country, and season.

Admission rates and 95% confidence intervals (CIs) per 100,000 person-years for RSV-coded and RSV-confirmed hospitalisations were calculated by age group, country and season using population data as of January 01 (Denmark, Finland, the Netherlands, and Spain-Valencia) or mid-year (England, Scotland). The 95% CIs were estimated using a Poisson exact method. In Spain-Valencia, the population was multiplied by the RSV circulation duration in years to account for seasonal differences in active monitoring periods (Supplementary Methods). In Scotland and England, no population data for the 2022/23 season was available, so the population was assumed to be as the 2021/22 population. An average (±SD) of hospital admission rates was calculated over the whole study period for each age group and country, and further stratified by COVID-19 period. To understand RSV-coding practices, we divided RSV-confirmed admission rates by the corresponding RSV-coded rates for each country, season and age group when enough sample size was available (≥10 patients).

Data collection was standardised using common data collection templates and definitions. Country-specific data challenges prevented the use of a general definition for some of the outcomes (detailed in Supplementary Methods). All analyses were performed in R (version 4.3.1) using common programming scripts to ensure comparability despite healthcare systems heterogeneity.

## Results

Table 1 shows the average (± SD) ≥18 years population among all seasons, together with the total number of RTI, and the total number and proportion of RSV-coded and RSV-confirmed admissions among adults ≥18 years during the whole study period.

**Table 1.** Average population and admission number among patients 18 years of age and above. The average ≥18 years population (± SD) during the whole study period is shown for each country. The total number of ≥18 years RTI admissions and the total number (and proportion from the RTI admissions) of RSV-coded and RSV-confirmed admissions are shown per country.

| Country | Study period | Average Population (± SD) | N RTI | N RSV-coded (%) | N RSV-confirmed (%) |
| --- | --- | --- | --- | --- | --- |
| Denmark | 2017/18-2021/22 | 4,652,883 ($\pm 47,104$) | 23,523 | 477 (0.2%) | 1,393 (0.6%) |
| England | 2017/18-2022/23 | 44,321,027 ($\pm 360,008$) | 4,305,867 | 21,587 (0.5%) | NA |
| Finland | 2016/17-2021/22 | 4,479,585 ($\pm 35,360$) | 337,751 | 5,078 (1.5%) | 6,259 (1.9%) |
| Netherlands | 2016/17-2020/21 | 13,951,809 ($\pm 182,643$) | 47,574 | 4,633 (1%) | NA |
| Scotland | 2016/17-2021/22 | 4,431,017 ($\pm 23,027$) | 272,105 | 1,185 (0.4%) | 2,057 (0.8%) |
| Spain-Valencia | 2016/17-2022/23* | 378,995 ($\pm 51,303$) | 8,752 | 48 (0.5%) | 442 (5.1%) |

\*Except 2020/21

The yearly average study population ranged from 378,995 (Spain-Valencia) to 44,321,027 (England). The total number of RTI admissions ranged from 8,752 (Spain-Valencia) to 4,305,867 (England). The proportion of RSV-coded admissions ranged from 1.5% (Finland) to 0.2% (Denmark). Among countries with available laboratory-confirmed data, RSV-confirmed proportions ranged from 5.1% (Spain-Valencia) to 0.6% (Denmark). Overall numbers and proportions stratified by COVID-19 periods are available in Supplementary Table S1. Numbers and proportions of admissions by age group, country, diagnosis group and COVID-19 stratification are available in Supplementary Table S2. Most of the RSV admissions were classified as Pneumonia (Supplementary Figure S1).

### RSV admission rates by season, country, and age group

RSV-coded hospitalisation rates per 100,000 person-years varied by country, age group and the COVID-19 period. For adults aged 18-64 years, average rates (± sd) ranged from 0.6 (± 0.2, Denmark) to 4.7 (± 3.4, Finland) before COVID-19 and from 1.1 (± 1.1, Spain-Valencia) to 4.8 (± 2.4, Finland) during COVID-19. For adults aged 65-74 years, rates ranged from 0 (± 0, Spain-Valencia) to 25 (± 23, Finland) before COVID-19 and from 5.1 (± NA, Denmark) to 25 (± 14, Finland) during COVID-19. Among those aged 75-84 years, rates were between 6 (± 2.7, Denmark) and 66 (± 64.3, Finland) before COVID-19 and between 13 (± NA, Denmark) and 45 (± 26, Finland) during COVID-19. For adults aged ≥85 years, rates varied from 7.3 (± 7.5, Spain-Valencia) to 166 (± 154, Finland) before COVID-19 and from 20 (± NA, Denmark) to 116 (± 64.4, Finland) during COVID-19 (Figure 1A, Supplementary Figure S2, Supplementary Figure S4A, Supplementary Table S3). The highest RSV-coded admission rates were observed in Finland, followed by England. Denmark and Spain-Valencia showed the lowest rates, likely due to data collection methods (ILI restriction in Spain-Valencia), RSV coding practices [26] or low number of diagnosis codes recorded at discharge in Spain-Valencia.

**Figure 1.**
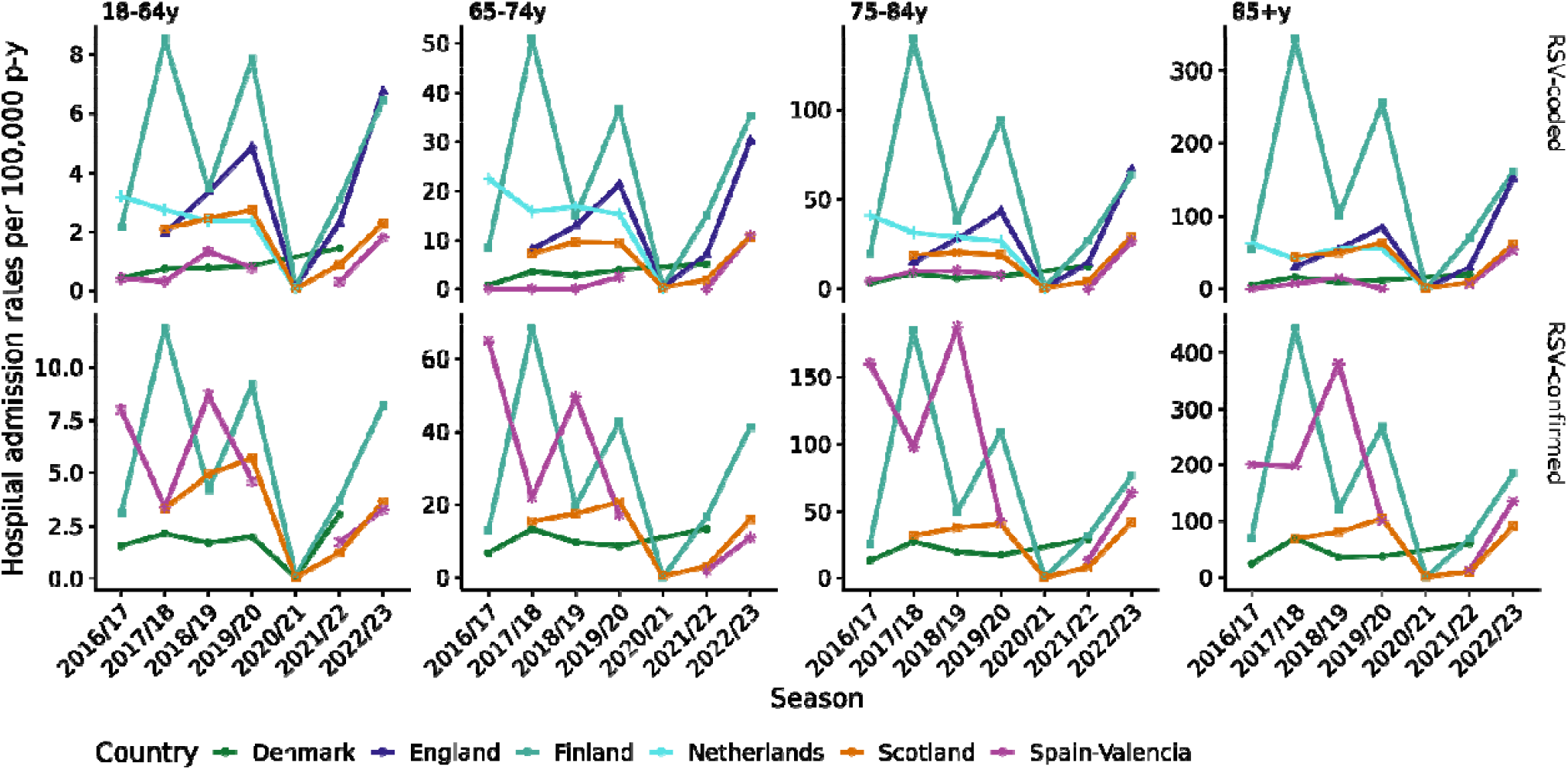
Hospitalisation admission rates (95%CI) in patients 18 years of age and above. The hospital admissions rates per 100,000 persons-year (y-axis) and season (x-axis) are shown for each country (colour), age group (vertical subpanels) and hospitalisation type: A) RSV-coded and B) RSV admissions with a laboratory-confirmed result for RSV. Note that each panel has a different y-axis scale to allow for readability. A summary figure with identical y-axis scales for all age groups can be found as Supplementary Figure S5.

RSV-confirmed admission rates per 100,000 person-years also varied substantially across countries, age groups and COVID-19 era. For adults aged 18-64 years, average lab-confirmed rates (± sd) ranged from 1.8 (± 0.3, Denmark) to 7 (± 2.9, Spain-Valencia) before COVID-19 and from 2 (± 1.7, Scotland) to 6 (± 3, Finland) during COVID-19. For adults aged 65-74 years, rates were between 10 (± 3, Denmark) and 45 (± 21, Spain-Valencia) before COVID-19 and between 6 (± 6, Spain-Valencia) and 29 (± 17, Finland) during COVID-19. For those aged 75-84 years, rates ranged from 20 (± 7, Denmark) to 148 (± 46, Spain-Valencia) before COVID-19 and from 25 (± 24, Scotland) to 54 (± 31, Finland) during COVID-19. Among adults aged ≥85 years, rates varied from 44 (±, 24, Denmark) to 260 (± 105, Spain-Valencia) before COVID-19 and from 51 (± 57, Scotland) to 127 (± 81, Finland) during COVID-19 (Figure 1B, Supplementary Figure S3, Supplementary Figure S4B, Supplementary Table S5). The highest RSV-confirmed admission rates were observed in Finland and Spain-Valencia. In Spain-Valencia, all included ILI patients were systematically tested for RSV, but in Finland, testing denominators were not known.

A drop in RSV admission rates was observed across all countries and age groups during the 2020/21 season. The incidence of RSV-coded or RSV-confirmed admissions ranged between 0 and 1.6 per 100,000 in 2020/21 considering all countries and age groups (Figure 1, Supplementary Figures S2-S3, and Supplementary Tables S3-S4, with rates returning to pre-pandemic levels from 2021/22 onwards. (Figure 1, Supplementary Tables S3-S4, and Supplementary Figures S2-S4). In England, a higher incidence of RSV-coded admissions was observed after 2020/21 (Supplementary Figure S4) across all age groups, but no laboratory-confirmed data were available to see whether the same trend was observed on RSV-confirmed hospitalisations. This trend was also observed in Denmark with both coded and lab-confirmed admissions without strong evidence, as only one season during COVID-19 was available and the error bars before COVID-19 overlap with the during COVID-19 bars (Supplementary Figure S4).

In some countries (Finland, Spain-Valencia), high RSV admission rates in a given year were often followed by lower rates the next year, as previously described in some EU countries (Finland, Norway and Denmark) [5]. This pattern was also seen across some age groups in Denmark before COVID-19 seasons (Figure S3). A high variability in rates was observed between countries, age groups and seasons, but rates consistently increased with increasing age regardless of the use of coded, lab-confirmed data, country or season (Supplementary Figure S5).

### RSV-coding practices

In countries with both RSV-coded and RSV-confirmed data, we estimated the degree of underestimation of RSV admission rates when relying solely on RSV codes. Denmark, Finland and Scotland had both types of data available and enough sample size (more than 10 RSV-coded admissions). In these countries, regardless of the age group and season, RSV-coded admissions resulted in an underestimation of the burden compared to RSV-confirmed admissions, except in 2021/22 in the ≥85 years age group in Finland (Figure 2 and Supplementary Figures S6-S7). In Finland, the average underestimation factor when relying on ICD-codes for admission rate estimations was 1.3 (± 0.1) times before COVID-19 and 1.2 (± 0.1) times during COVID-19. In Scotland, the average underestimation factor was 1.8 (± 0.2) times before COVID-19 and 1.5 (± 0.2) times during COVID-19. Denmark showed the highest underestimation factor, with an average underestimation factor of 3.5 (±0.6) times before COVID-19 and 2.5 (± 0.4) times during COVID-19. A figure including admissions of less than 10 patients in the ICD-coded dataset is available as Supplementary Figure S7.

**Figure 2.**
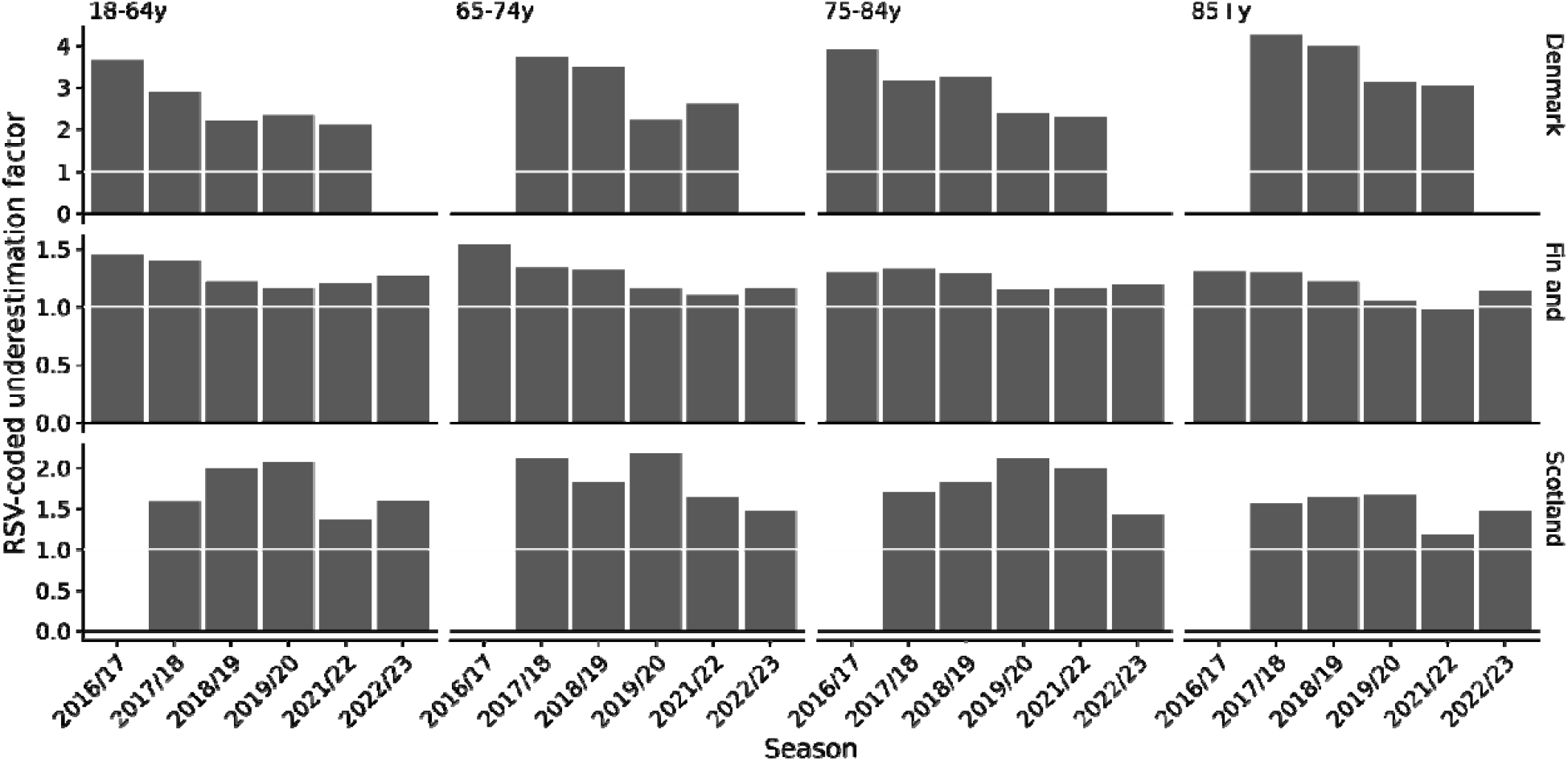
RSV-coding practices and its underestimation. The RSV-coded underestimation factor (y-axis) has been obtained by dividing in each season (x-axis), country (vertical subpanels) and age group (horizontal subpanels) the corresponding RSV-confirmed admission rate by the corresponding RSV-coded admission rate. Numbers above the white line indicate that the incidence estimated with RSV-coded admissions is lower than the one estimated with RSV-confirmed. Groups with less than 10 patients in the RSV-coded dataset have been removed in this Figure. A complete picture including >10 patient′s group is available in Supplementary Figure S7.

### Length of stay of RSV-associated hospitalisations

Figure 3 displays the median length of stay (LOS) for each hospitalisation type, age group, season and country. The median LOS for an RSV admission (RSV-coded or confirmed) with enough sample size (>10 patients) ranged between 2.0 days (2017/18, Scotland) to 8.0 days (2017/18, Spain-Valencia) in the 18-64 years age group, between 2.0 days (2017/18, Scotland) to 8.0 days (2016/17, Netherlands) in the 65-74 year age group, 1 day (2019/20, Scotland) to 12.5 days (2020/21, England) in the 75-84 years age group and 1 day (2019/20, Scotland) to 9.5 (2021/22, England) in the ≥85 years age group. The lowest LOS were always evident within each age group, except in Scotland and England, where a longer LOS was noted in some age groups during 2020/21 for both RSV-coded and RSV-confirmed admissions, but these were not statistically significant (Figure 3). The variability between seasons was higher for RSV-coded/confirmed hospitalisations compared to RTI admissions, but the number of admissions used for the LOS estimations was higher for RTI admissions. RSV-related hospitalisations did not result in longer stays compared to general RTI admissions (Supplementary Figure S8).

**Figure 3.**
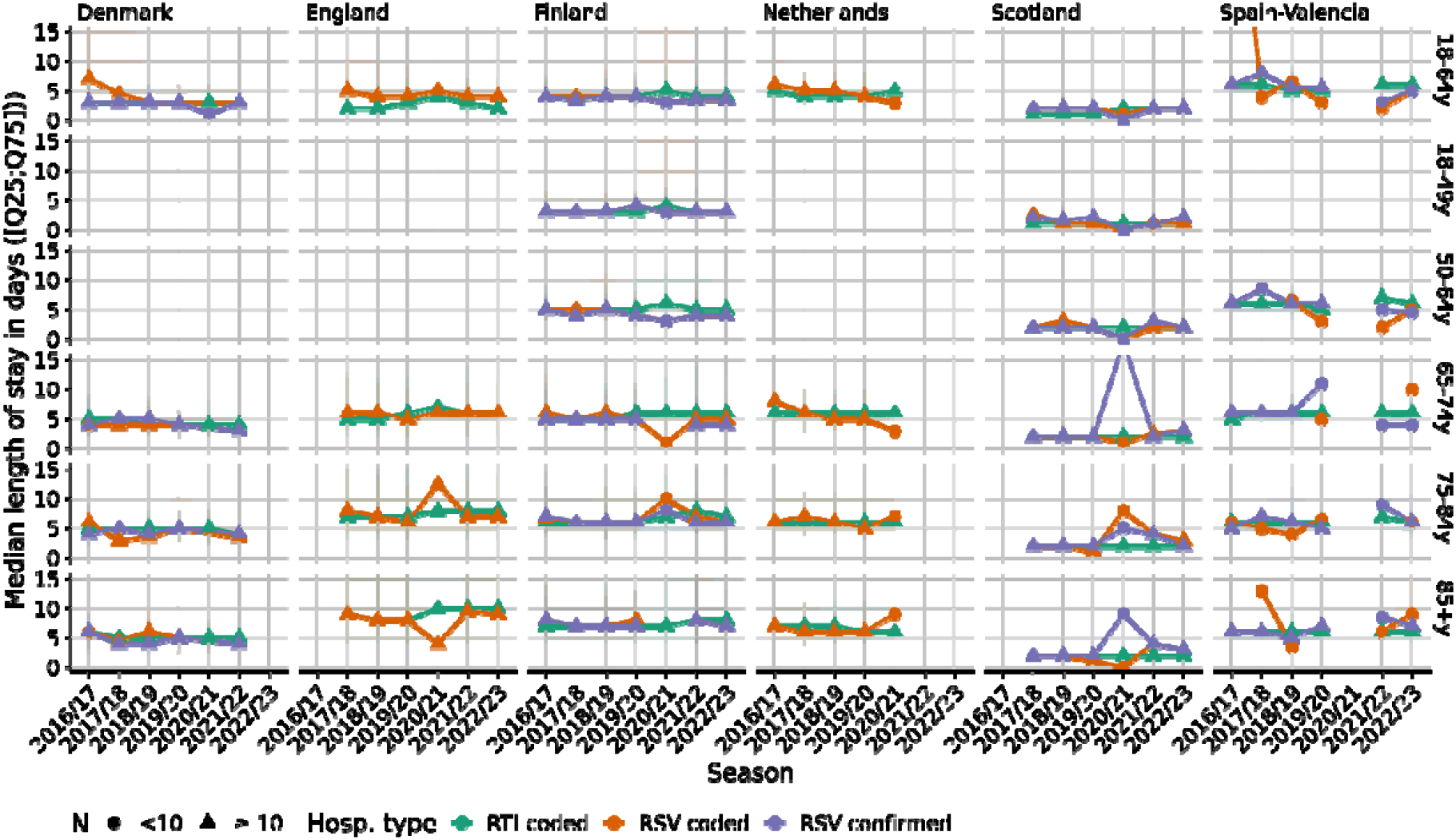
Hospitalisations median length of stay. Median LOS (y-axis) and 25/75 quartiles (shadow area) of all hospital admissions per country, age group and seasons (x-axis). Each hospitalisation type is shown with a different colour (green: RTI admissions, orange: RSV-coded and violet – RSV-confirmed admissions). If the estimate is based on less than 10 patients the shape is represented as a dot, whereas if the estimate is based on more than 10 patients the shape is represented as a triangle. Outliers (18-64 age group and RSV-coded in Spain-Valencia: 52 days [1 patient] and 65-74 age group and RSV-confirmed in Scotland [2 patients]: 18 days) are not shown to allow for readability of the graph.

### RSV-associated ICU-admissions

In England, ICU admissions were available and are summarised in Table 2. Across all age groups, a higher ICU admission proportion among RSV-coded patients was observed before the COVID-19 pandemic The ICU admission proportion decreased with increasing age both before and during COVID-19. Before COVID-19, in the 18-64- and 65-74-years age groups, the RSV-coded ICU admission proportions were higher than the overall RTI admission proportions (7.2% versus 5.8% and 7.9% versus 6.1%, respectively). This trend has not been observed since COVID-19. Numbers stratified by diagnosis group are available in Supplementary Table S4.

**Table 2.** Average (±SD) RTI and RSV-associated ICU admissions in England of the seasons before COVID-19 (2016/17-2018/19) and of the seasons during COVID-19 (2021/22-2022/23) stratified by age group. The proportion of RTI/RSV-coded ICU admissions to the total number of RTI or RSV-coded admissions is also shown.

| Age group | COVID-19 | Avg. (±SD) RTI-ICU admissions, % | Avg. (±SD) N RSV-coded ICU admissions, % |
| --- | --- | --- | --- |
| 18-64 | Before | 12468 (±292), 5.8% | 65 (±23), 7.2% |
| | During | 10318 (±5387), 3.8% | 47 (±7), 3.0% |
| 65-74 | Before | 7372 (±332), 6.1% | 46 (±7), 7.9% |
| | During | 4939 (±2275), 3.8% | 22 (±3), 2.1% |
| 75-84 | Before | 6338 (±193), 3.7% | 27 (±11), 3.9% |
| | During | 3716 (±1380), 1.9% | 12 (±4), 0.9% |
| ≥85 | Before | 1996 (±97), 1.2% | 5 (±0), 0.9% |
| | During | 882 (±254), 0.5% | 5 (±1), 0.4% |

### RSV-associated case fatality rates (CFR)

In countries with available mortality data (England, Finland and Spain-Valencia), the CFR among RSV-coded hospitalisations ranged from 0-3.3% before COVID-19 and 0-3.5% during COVID-19 for the 18-64 years age group, 0-7.4% before COVID-19 and 0-8.2% during COVID-19 for the 65-74 years age group, 5.7-12.9% before COVID-19 and 0-10.8% during COVID-19 for the 75-84 years age group, and 11.9-30% before COVID-19 and 6.2-17.6% during COVID-19 for the ≥85 years and older age group (Table 3). Among RSV-confirmed hospitalisations (Finland and Spain-Valencia, respectively), the CFR was 2.5 and 0% before COVID-19, and 1 and 0% during COVID-19 for the 18-64 years age group, 3.5 and 5.3% before COVID-19 and 2.2 and 0% during COVID-19 for the 65-74 years age group, 6.5 and 2.4% before COVID-19 and 3.4 and 0% during COVID-19 for the 75-84 years age group, and 12 and 11% before COVID-19 and 6 and 15% during COVID-19 for the ≥85 years age group (Table 3). Numbers stratified by diagnosis group are available in Supplementary Table S5.

**Table 3.** Average (±SD) RTI and RSV-associated admissions that end up in death in England, Finland and Spain-Valencia of the seasons before COVID-19 (2016/17-2018/19) and of the seasons during COVID-19 (2021/22-2022/23) stratified by age group. The CFR to the total number of RTI or RSV-coded/confirmed admissions is also shown.

| Age group | COVID-19 | England |  | Finland |  |  | Spain-Valencia |  |  |
| --- | --- | --- | --- | --- | --- | --- | --- | --- | --- |
|  |  | Avg. N RTI deaths, CFR | Avg. N RSV-coded deaths, CFR | Avg. N RTI deaths, CFR | Avg. N RSV-coded deaths, CFR | Avg. N RSV-confirmed deaths, CFR | Avg. N RTI deaths, CFR | Avg. N RSV-coded deaths, CFR | Avg. N RSV-confirmed deaths, CFR |
| 18-64 | Before | 8391 (±232), 3.9% | 30 (±1), 3.3% | 481 (±25), 3.7% | 4 (±3), 2.8% | 5 (±4), 2.5% | 6 (±3), 1.2% | 0 (±0), 0% | 0 (±0), 0% |
| | During | 10550 (±1368), 3.9% | 54 (±42), 3.5% | 402 (±227), 3.5% | 1 (±1), 0.6% | 2 (±0), 1% | 2 (±0), 0.7% | 0 (±0), 0% | 0 (±0), 0% |
| 65-74 | Before | 12660 (±612), 10.4% | 43 (±3), 7.4% | 995 (±28), 8.6% | 7 (±8), 4% | 8 (±10), 3.5% | 13 (±2), 3.3% | 0 (±0), 0% | 1 (±2), 5.3% |
| | During | 14282 (±1039), 10.9% | 86 (±71), 8.2% | 958 (±408), 9.2% | 4 (±2), 2% | 4 (±4), 2.2% | 4 (±2), 2.5% | 0 (±0), 0% | 0 (±0), 0% |
| 75-84 | Before | 24388 (±1705), 14.1% | 54 (±14), 7.8% | 1910 (±143), 12.2% | 13 (±14), 5.7% | 20 (±23), 6.5% | 27 (±9), 5.0% | 0.3 (±0.7), 12.9% | 1 (±1), 2.4% |
| | During | 28196 (±26), 14.6% | 152 (±148), 10.8% | 1736 (±687), 11.5% | 7 (±7), 3.5% | 8 (±8), 3.4% | 9 (±4), 4.1% | 0 (±0), 0% | 0 (±0), 0% |
| ≥85 | Before | 34054 (±3645), 19.8% | 69 (±20), 11.9% | 2559 (±163), 17.8% | 30 (±27), 12.3% | 38 (±42), 12.3% | 48 (±22), 9.7% | 0.3 (±0.7), 30% | 4 (±1), 11.12% |
| | During | 35145 (±878), 19.7% | 219 (±238), 17.6% | 2110 (±797), 16.6% | 12 (±9), 6.2% | 12 (±11), 6.2% | 12 (±4), 5.6% | 0.5 (±0.5), 12.5% | 2 (±2), 15% |

## Discussion

This study highlights the complex epidemiology of RSV in adults across several European countries, particularly in the context of the COVID-19 pandemic. The comparison of RSV-coded versus RSV-confirmed hospitalisations reveals significant discrepancies, highlighting challenges in capturing the true burden of RSV-related illnesses. Our data, encompassing multiple countries with varied healthcare systems, provide valuable insights into RSV epidemiological trends. Notably, the study reveals consistent patterns of RSV circulation and age-specific hospitalisation rates despite differing national health practices, suggesting a broader applicability of the findings. This consistency is particularly relevant given the varying public health measures in place during the study period, including NPIs aimed at curbing COVID-19.

Across all countries the highest hospitalisation rate was observed in adults aged ≥85 years with up to 445 laboratory-confirmed hospitalisations per 100,000 person-years. During 2020/21, when the strongest COVID-19 NPI measures were in place, almost no RSV-associated hospitalisations occurred. No significant differences in the LOS of RSV-associated hospitalisations compared to all-cause RTI admissions were observed. Both RSV-associated ICU-admissions and RSV CFRs showed no significant differences compared to RTI ICU-admissions and RTI CFRs. RSV and RTI CFRs increased with increasing age and ICU-admissions decreased with increasing age (Table 2 and Table 3).

Country-specific nuances were observed with England, Finland, the Netherlands, and Scotland displaying similar RSV-coded admission trends. This likely reflects similarities in healthcare systems and coding practices. Biennial peaks of RSV admissions were noted in Finland, Spain-Valencia and Denmark, consistent with previous observations [27]. Higher incidences of RSV-confirmed admissions were seen in countries with robust access to laboratory data (Finland and Spain-Valencia, in the latter systematic PCR testing was performed in all included patients). Yet, it is worth noting that diagnostic tests and systematic testing there still may be an underestimation of at least 2.2 times [28, 29].

In routinely collected healthcare records, access to laboratory test results is often not possible, and even when available, the testing denominators are commonly not provided. Thus, RSV-coding practices become the best available proxy for estimating RSV burden. A study in Denmark showed that ICD-coded admissions underestimate the true RSV burden [26]. Here, we also observed that relying on RSV codes led to underestimation factors between 1.1 times (Finland) and 4.3 times (Denmark). Finland showed a high correlation between RSV-coded and RSV-confirmed admissions (Pearson r^2^ = 0.99), reflecting high quality clinical practices. Yet, similar patterns between RSV-coded and RSV-confirmed admissions were observed in Scotland, Finland, and Denmark, suggesting that, despite its limitations, RSV-coded admissions could be used as an indicator of RSV epidemiology when laboratory data are unavailable [27].

RSV epidemiology in older adults appears to be stabilising to pre-COVID-19 patterns, contrasting with the increases observed in paediatric populations [18, 30, 31], due to an immunity debt hypothesis [32] /an increase in RSV testing [33]. In England, a higher rate of RSV-coded admissions was observed during COVID-19, but only two seasons data was used and the trend was not significant (Supplementary Figure S8). Our data included up to three seasons before COVID-19 and two during-COVID-19, which included season 2021/22, where NPI measures were still in place in most countries [34]. This may have impacted the estimates. Future data on RSV-related hospitalisations in older adults are needed to fully understand during-COVID-19 trends.

The data collection method in Spain-Valencia differed as it relied on an active surveillance network setup for influenza surveillance and on the ILI case definition, which is well-known to underestimate the real RSV burden [11, 14, 35]. Moreover, data in each season was adjusted to the RSV circulation period and the admission rates were shown per 100,000 adults during the RSV circulation period. Despite this, similar trends in seasonality, rates, and age distribution of RSV-confirmed cases were observed between Finland and Spain-Valencia (RSV-confirmed datasets Pearson r^2^ =0.75). This suggests that routinely collected national healthcare data can still provide valuable insights into RSV trends and epidemiology in older adults despite limitations.

Our study presents several limitations that must be considered when interpreting the findings. The reliance on routinely collected national healthcare data introduces potential biases, including coding inaccuracies and inconsistencies in clinical practices across different countries. Particularly, the ICD-coded admissions likely underestimate the true burden of RSV, as evidenced by underestimation factors ranging from 1.1 times in Finland to 4.3 times in Denmark. The disparity in coding practices, especially during COVID-19, and the incomplete understanding of RSV testing practices further complicates the analysis and interpretations. The variability in data collection methods and the limited scope of laboratory-confirmed cases restrict direct comparability across countries. These limitations underscore the need for more systematic and standardised testing approaches to improve the accuracy and reliability of RSV burden estimates. The use of other approaches for RSV-burden estimation such as time series modelling [36] could help in overcoming some of these limitations.

Despite these limitations, this study provides a comprehensive overview of RSV-associated hospitalisations in adults across multiple European countries, offering critical insights into RSV epidemiology in the context of the COVID-19 pandemic. The observed stabilisation of RSV patterns in older adults post-COVID-19 contrasts with increases seen in paediatric populations, raising questions about age-specific immunity and the impact of NPIs. Despite the inherent limitations related to coding practices and variability in data collection methods, the observed trends are indicative of the significant public health burden posed by RSV in older adults. The introduction of new RSV immunisation strategies underscores the necessity for reliable epidemiological data to guide public health policies and interventions. As such, this study lays the groundwork for future research and surveillance efforts aimed at mitigating the impact of RSV, particularly in the context of evolving viral dynamics and public health landscapes.

## Supporting information

Suppementary Fig 1

Suppementary Fig 2

Suppementary Fig 3

Suppementary Fig 4

Suppementary Fig 5

Suppementary Fig 6

Suppementary Fig 7

Suppementary Fig 8

## Ethical statement

All countries participating in this study received ethical and/or institutional board approvals to access and utilise national and/or regional health registries.

## Data availability

All data are presented in this manuscript. Written requests may be made to the relevant data controllers to access specific national or regional data.

## Acknowledgements

We thank the peripheral diagnostic laboratories spread throughout the Netherlands and the Health iQ Limited, trading as CorEvitas, which has a data sharing agreement in place with NHS Digital to store a copy of the latest 5 years of Hospital Episode Statistics data in-house and provided England data. Authorised Health iQ employees accessed to this in-house database to identify which the study population and conduct data aggregation. Copyright © (2023), re-used with the permission of The Health & Social Care Information Centre. All rights reserved.

We acknowledge the support of the electronic data research and innovation services (eDRIS) team at Public Health Scotland for their involvement in obtaining approvals, provisioning and linking, and the use of the secure analytical platform with the National Safe Haven.

## Authors’ contributions

OJ, AUF, CKJ, TL, DG, RAC, RK, TH, TKF, MvB, ROY, HN, and HC contributed to the design of the study. OJ, AUF, CKJ, TL, MvB, RAC, and ROY contributed to the data extraction for this work and contributed to the analysis of the data, OJ and AUF contributed to the data visualization and statistical analyses, AUF and ROY wrote the manuscript, all authors reviewed and validated this manuscript.

