## Supplementary file for "RSV healthcare burden in adults before and since the emergence of the COVID-19 pandemic in 6 European countries"

**Supplementary Materials**

**Methods**

1. **National registries**

**Denmark**

The Danish data is based on the Danish National Patient Registry (NPR) [1-4]. The NPR provides nationwide longitudinal registration of detailed administrative and clinical data [1-4]. For each patient contact, one primary and numerous, optional, secondary diagnoses are recorded. For this study, data on all admissions with an RSV diagnosis were gathered from 2016 to 2023. Linkage to RSV-positive laboratory confirmations was possible through data from the national clinical microbiological database, where data from all microbiological tests are stored. This data was provided by Statens Serum Institute through the KIDS database. Patients identified in DNPR were also linked to the Medical Birth Registry (MBR), as well as the Civil Registration System (CRS) to obtain additional and missing information. The MBR contains data on all births in Denmark since 1973 including background information on the circumstances of the birth. No information on SARS-CoV-2 was available. When the number of admissions or deaths was between 1 and 4, information was provided as “<5” and treated in the analysis as 2 to enable display on one hand and aggregation over multiple values of the other hand. Demographic data for population estimates were collected from Statistics Denmark [5].

**England**

Hospitalisation and death data were collected from the England Hospital Episode Statistics (HES) [6] database that monitors >98% of England’s population and gathers information related to the patient and their clinical course in the hospital. HES is a reimbursement dataset that facilitates secondary care provider reimbursement from local health commissioners. HES contains details of inpatient, including ICU, admissions in a dataset called Admitted Patient Care (APC), from National Health Service (NHS) hospitals in England. Patient secondary care activity is longitudinally linked across all NHS hospitals. The information within HES is collected across services as part of the Commissioning Data Sets. Data are submitted monthly to NHS Digital for processing and is made available as the Secondary Uses Service dataset which can be used for non-clinical purposes, such as research and planning health services. In HES, APC diagnoses are recorded using the ICD-10 and procedures performed using the Office of Population, Censuses and Surveys Classification of Surgical Operations and Procedures, fourth revision (OPCS-4). Inpatient admissions may have up to 20 ICD-10 diagnoses, of which one is the primary cause of admission, investigation, or treatment, and the remaining 19 codes are secondary diagnoses codes. The same is available for procedures with up to 20 OPCS-4 codes. ICU episodes can be level two (high dependency) and level three (intensive organ support and/or nursing care). ICU episodes can occur multiple times during the same overnight admission as patient care is escalated and de-escalated. When a patient dies during a hospital stay, this is recorded in the HES APC dataset as an inpatient death. However, deaths that occur among patients outside of the inpatient setting are not recorded in this dataset. To exclude scheduled admissions, the HES admission method field was used, and all hospitalisations that were labelled as “Waiting list”, “Booked”, or “Planned” were excluded. The exclusion criterion for admissions shorter than 12 hours was not applied, as the admission duration in HES can only be calculated in whole days. Admissions coded as COVID-19 that occurred prior to 01 March 2020 were excluded, as these were likely to be coding errors. Where the age was missing, it was imputed using information from any other admissions for that person where the age was recorded. From this, an approximate date of birth was calculated and then used to calculate an approximate age at the time of the admission with the missing age. When nothing was available to impute age, the admission was counted in the “Overall” age group category only. The earliest occurrence of each risk factor was identified for each person, using all HES admissions, including non-RTI admissions. People were considered to have that risk factor from that point forward. Any admission occurring at or after the risk factor was first observed was counted under that risk factor. The population comes from various United Kingdom censuses from the National Office for Statistics: for 2021/2022 the population described in Census 2021 [7], for previous seasons’ populations the mid-year census of the year, i.e., week 27 of the season [8]. For 2022/2023, as the census was not yet publicly available at the time of the study, we assumed the same population for mid-2022 as the one in mid-2021 to compute the incidence, which is expected to have low bias on the outcomes as the population is not varying a lot over years.

**Finland**

The Finnish data is based primarily on two nationwide registers maintained by the Finnish Institute for Health and Welfare (THL): the Finnish Care Register for Health Care (HILMO) and the National Infectious Diseases Register (NIDR). HILMO covers individual level clinical and administrative data from inpatient care, specialised outpatient care and day surgeries from both hospitals and other institutions. Each registered event has a symptom-cause pair of primary diagnoses and similar pairings of optional secondary diagnoses, all of which have been recorded using the ICD-10 classification since 1996. For this study the data from HILMO was gathered from 2016 to 2023. The NIDR captures records of selected microbiological findings and related diagnoses that all laboratories and physicians are obliged to report under the Communicable Disease Act. All positive test results for RSV, influenza A virus and influenza B virus are included in the register, though there is no denominator information on the total number tested. General demographic records and dates of death for the study population were obtained from the Population Information System which is maintained by Statistics Finland. Risk group variables for infants were collected from Medical Birth Register (MBR). The MBR dataset covers only the years 2018-2020. The linkage of data from different registers was carried out using unique personal identification codes.

**Scotland**

In Scotland, we used the Scottish Morbidity Record (SMR01) and Electronic Communication of Surveillance in Scotland (ECOSS) registries containing individual-level patient data on all inpatient and day cases in hospitals and laboratory cases. We used the continuous inpatient stay (CIS) marker to group admissions belonging to the same episode; for example, transfers from one hospital or significant facility to another were included as the same admission. The period of data availability was between January 2017 to June 2023, and we only integrated cases within the defined surveillance seasons (week 27 of year Y to week 26 of year Y+1). The study was approved by the Public Benefit and Privacy Panel for Health and Social Care (HSC-PBPP), and data were provided by the electronic Data Research and Innovation Services (eDRIS) [9, 10].

**Netherlands**

The Dutch Hospital Data (DHD) registration collects, manages and processes hospital data and manages standards for its registration. The used data sources were LBZBASISTAB and LBZDIAGNOSENTAB for the years 2016 to 2021. All diagnoses were recorded using ICD-10 classification. The demographic database from Statistics Netherlands (CBS) GBAPERSOONTAB was used to link data on gender and birth date to the admitted patients. Linkage to RSV-positive confirmations was not possible for this study. This non-public microdata was provided by Statistics Netherlands [11].

**Spain-Valencia**

In Spain-Valencia, patient information was collected through the Valencia Hospital Surveillance Network for the Study of influenza and other Respiratory Viruses (VAHNSI), an active prospective hospital-based surveillance network (~1 million catchment population; 21% of the total Valencia population). The network is coordinated by the Vaccine Research Department of FISABIO-Public Health and has been described previously [12-15]. Patients had to fulfil the following criteria to be included in the study: being hospitalised via emergency room with a diagnosis compatible with an RTI, residing in the catchment area of one of the participating hospitals for at least 6 months, non-institutionalised, not discharged from a previous hospital admission in the last 30 days and give their (or their legally authorised representative) written consent. Patients ≥ 18 years old were included if, upon admission, they met symptoms compatible with the Influenza-like-Illness case definition, defined as the presence of at least one respiratory symptom (cough, sore throat or shortness of breath) with an onset within 7 days prior to admission [16]. As VAHNSI is a surveillance network initially set up to cover influenza seasons, monitoring did not occur throughout the whole year and the duration of the monitoring was also different across seasons. Therefore, data was adjusted to the RSV circulation period in each of the season to allow data comparison across years with different surveillance lengths. Circulation was defined as the weeks between the first of at least two consecutive weeks with two or more RSV cases and the week before the first of at least two consecutive weeks without RSV cases considering the PCR results of included patients from all ages. The loss of RSV confirmed cases after adjusting the data to the circulation period seasons is negligible. The duration in weeks of each season was calculated as the total number of epidemiological weeks in each RSV circulation period. The population was adjusted to the length of the RSV circulation period in years (approximated as 1 year; ~ 52.143 weeks). Table 1 and 2 describe the hospitals involved and the RSV circulation period in each season. Due to the COVID-19 pandemic no data was collected during season 2020/2021. During 2021/22 there were two RSV circulation periods (from W43 to W05 and from W13 to W26) and data from both circulations have been used in the study.

**Table 1** Participating hospitals and their overall catchment size

| **Hospital** | **Season** | **Catchment size** |
| --- | --- | --- |
| General Castellón | All | 279,111 |
| La Fe | All | 284,152 |
| Dr. Peset | All | 272,842 |
| General Alicante | Until 2019/20 | 271,120 |
| La Marina Baixa | 2021/22 and 2022/23 | 184,076 |

**Table 2** Data collection period per season in Spain-Valencia

| **Season** | **RSV-circulation starts** | **RSV-circulation ends** | **N weeks** | **Length (years)** |
| --- | --- | --- | --- | --- |
| 2016/17 | 2016-W45 | 2017-W11 | 18 | 0.34 |
| 2017/18 | 2017-W44 | 2018-W17 | 25 | 0.48 |
| 2018/19 | 2018-W44 | 2019-W14 | 22 | 0.42 |
| 2019/20 | 2019-W44 | 2020-W11 | 19 | 0.36 |
| 2021/22** | Circ1: 2021-W43  Circ2: 2022-W13 | Circ1: 2022-W05  Circ2: 2022-W26 | 28 | 0.54 |
| 2022/23 | 2022-42 | 2023-12 | 22 | 0.42 |

** Two RSV circulation periods

No follow-up of patients was done after hospital discharge.

The study protocol was approved by the Ethics Research Committee of the Dirección General de Salud Pública-Centro Superior de Investigación en Salud Pública (DGSP-CSISP). All subjects signed written informed consent prior to their inclusion in the study.

2. **Country-specific data availability**

Table 3 summarises the data availability of each of the participating countries

**Table 3** Data availability per country.

| **Data availability** | **Scotland** | **England** | **Netherlands** | **Finland** | **Denmark** | **Spain-Valencia** |
| --- | --- | --- | --- | --- | --- | --- |
| **Population coverage** | National | National | National | National | National | ~21% Spain-Valencia region |
| **Denominator population of ≥18-year-olds**  **[Mean (range) over study period]** | 4,431,017 (4,394,745,  4,454,919) | 44,321,027 (43,752,473,  44,715,443) | 13,951,809 (13,701,285,  14,190,874) | 4,479,585  (4,431,392,  4,537,778) | 4,652,883  (4,580,547,  4,721,691) | 378,995  (306,076,  456,659) |
| **Years available for this study** | 2017-2022 | 2017-2023 | 2016-2021 | 2016-2023 | 2016-2022 | 2016-2023 (except 2020/21) |
| **Linkage to RSV laboratory confirmations** | Yes | No | No | Yes | Yes | Yes |
| **Coding system** | ICD10 | ICD10 | ICD10 | ICD10 | ICD10 | ICD-9, ICD10 |
| **Maximum number of diagnosis codes per record** | 6 | 20 | No maximum | No maximum | No maximum | 3 |
| **Mortality data** | No | Yes | No | Yes | Yes | Yes |
| **ICU admissions** | No | Yes | No | No | No | No |
| **Risk group**  **identification** | No | Yes | No | Yes | Yes | Yes |

3. **List of ICD codes**

In Table 4, the list of ICD-10 and ICD-9 codes used to identify RTI and RSV-coded (in red) hospitalisations. The codes are shown together with their corresponding diagnosis group classification.

**Table 4** List of ICD-10 and ICD-9 codes used in this study, with RTI codes in black and RSV codes in red.

| **Diagnosis classification** | **ICD-10** | **ICD-9** |
| --- | --- | --- |
| **Acute upper respiratory tract infection (URTI)** | J00 J02.0 J02.8 J02.9 J03.0 J03.8 J03.9 J04.0 J04.1 J04.2 J05.0 J05.1 J06.0 J06.8 J06.9 | 460 4610 4611 4612 4613 4618 4619 462 463 46400 46401 46410 46411 46420 46421 46430 46431 4644 46450 46451 4650 4658 4659 |
| **Pneumonia & Influenza (LRTI)** | J09 J10.0 J10.1 J10.8 J11.0 J11.1 J11.8 J12.0 **J12.1** J12.2 J12.3 J12.8 J12.9 J13 J14 J15.0 J15.1 J15.2 J15.3 J15.4 J15.5 J15.6 J15.7 J15.8 J15.9 J16.0 J16.8 J17.0 J17.1 J17.2 J17.3 J17.8 J18.0 J18.1 J18.2 J18.8 J18.9 | 4800 **4801** 4802 4803 4808 4809 481 4820 4821 4822 48230 48231 48232 48239 48240 48241 48249 48281 48282 48283 48284 48289 4829 4830 4831 4838 4841 4843 4845 4846 4847 4848 485 486 4870 4871 4878 514 |
| **Bronchiolitis & Bronchitis (LRTI)** | J20.0 J20.1 J20.2 J20.3 J20.4 **J20.5** J20.6 J20.7 J20.8 J20.9 **J21.0** J21.1 J21.8 J21.9 J40 | 4660, **46611**, 46619 |
| **Unspecified LRTI** | J22 | 5198 (not exact map) |
| **SARS-CoV-2/COVID-19** | U07.1, U07.2, U08-10 | - |
| **RSV-specific codes** | **B97.4** | **0796** |

**Results**

**Table S1 Average population and number of RTI, RSV-coded and RSV-confirmed admissions among patients 18 years of age or older stratified by before and during COVID-19.** The average population (and standard deviation) for patients ≥18 years of age is shown for each country and COVID-19 period (before: 2016/17-2018/19 seasons and during: 2021/22-2022/23 seasons, depending on the country). The total number of RTI admissions analysed per country is shown for each COVID-19 period. The number of RSV-coded and RSV-confirmed admissions, together with the proportion from the RTI admissions is also shown.

| **Country** | **Average Population**  **(±SD)** | | **N RTI** | | **N RSV-coded**  **(%)** | | **N RSV-confirmed**  **(%)** | |
| --- | --- | --- | --- | --- | --- | --- | --- | --- |
|  | **before** | **during** | **before** | **during** | **before** | **during** | **before** | **during** |
| **Denmark** | 4,613,978  (±27,809) | 4,721,691  (±NA) | 133,485 | 36,223 | 214,  0.2% | 165,  0.5% | 751,  0.6% | 400,  1.1% |
| **England** | 43,887,516  (±144,368) | 44,715,443 (±0) | 1,365,529 | 1,548,738 | 5,510, 0.4% | 10,48,  0.7% | NA | NA |
| **Finland** | 4,446,030  (±12,141) | 4,525,251  (±13,392) | 164,193 | 99,614 | 2,395, 1.5% | 1,423,  1.4% | 3,178,  1.9% | 1,663, 1.7% |
| **Netherlands** | 13,823,706  (±106,128) | NA | 319,414 | NA | 3,555, 1.1% | NA | NA | NA |
| **Scotland** | 4,401,524  (±7,247) | 4,454,919  (±0) | 109,555 | 7,561 | 494, 0.5% | 390,  0.5% | 880,  0.8% | 584,  0.8% |
| **Spain-Valencia** | 371,827  (±50,557) | 410,329  (±49,529) | 5,719 | 1,837 | 16,  0.3% | 27,  1.5% | 335,  5.9% | 65,  3.5% |

**Table S2 Numbers and proportions admissions.** The average (±SD) number of RTI, RSV-coded and RSV-confirmed admissions is shown for each age group, country, diagnosis group and COVID-19 period (before: 2016/17-2018/19 seasons and during: 2021/22-2022/23 seasons, depending on the country). The proportion from the RTI admissions for the RSV-coded and RSV-confirmed admissions is also shown.

|  |  | **Denmark** | | | **England** | | **Finland** | | | **Netherlands** | | **Scotland** | | | **Spain-Valencia** | | |
| --- | --- | --- | --- | --- | --- | --- | --- | --- | --- | --- | --- | --- | --- | --- | --- | --- | --- |
| **Age group** | **COVID-19** | **RTI** | **RSV-coded** | **RSV-confirmed** | **RTI** | **RSV-coded** | **RTI** | **RSV-coded** | **RSV-confirmed** | **RTI** | **RSV-coded** | **RTI** | **RSV-coded** | **RSV-confirmed** | **RTI** | **RSV-coded** | **RSV-confirmed** |
| 18-64 | Before | 11467(10534,12400) | 23(16,30), 0.2% | 63(53,73), 0.5% | 216660 (207718, 225602) | 898 (556,1240), 0.4% | 13155 (12521,13789) | 154 (44, 264), 1.2% | 210 (54,366), 1.6% | 26896 (20341,33451) | 293 (250,336), 1.1% | 17600 (17354,17846) | 77 (69,85), 0.4% | 140 (103,177), 0.8% | 464 (351,577) | 2 (0,4), 0.4% | 19 (11,27), 4.1% |
|  | During | 8237 (±NA) | 51(±NA), 0.6% | 108(±NA), 1.3% | 271297 (245283 ,297311) | 1552 (474,2630), 0.6% | 11576 (10608,12544) | 154 (77, 231), 1.3% | 194 (91,297), 1.7% | NA | NA | 11894 (8124,15664) | 54 (21,87), 0.5% | 82 (24,140), 0.7% | 298 (231,365) | 3 (0,6), 1% | 8 (6,10), 2.7% |
| 65-74 | Before | 10443(9808,11078) | 15(6,24), 0.1% | 64(43,85), 0.6% | 121223 (119079,123367) | 582 (392,772), 0.5% | 11583 (10883,12283) | 168 (13, 323), 1.5% | 228 (22,434), 2% | 28969 (22315,35623) | 343 (282,404), 1.2% | 10442 (9796,11088) | 48 (38,58), 0.5% | 94 (85,103), 0.9% | 401 (327,475) | 0, 0% | 19 (12,26), 4.7% |
|  | During | 7645 (±NA) | 32(±NA), 0.4% | 84(±NA), 1.1% | 131280 (128150,134410) | 1038 (123,1953), 0.8% | 10410 (9312,11508) | 176 (78, 274), 1.7% | 202 (82,322), 1.9% | NA | NA | 7279 (5916,8642) | 38 (1,75), 0.5% | 56 (2,110), 0.8% | 182 (175,189) | 2 (-2,6), 1.1% | 3 (0,6), 1.6% |
| 75-84 | Before | 12814(12232,13396) | 21(11,31), 0.2% | 70(44,96), 0.5% | 172590 (169289,175891) | 694 (382,1006), 0.4% | 15609 (14614,16604) | 235 (7, 463), 1.5% | 310 (5,615), 2% | 31120 (23038,39202) | 338 (283,393), 1.1% | 14224 (12801,15647) | 64 (59,69), 0.4% | 114 (99,129), 0.8% | 544 (452,636) | 2 (1,3), 0.4% | 41 (29,53), 7.5% |
|  | During | 11830(±NA) | 56 (±NA), 0.5% | 129(±NA), 1.1% | 193230 (179031,207429) | 1406 (119,2693), 0.7% | 15102 (12262,17942) | 198 (78, 318), 1.3% | 234 (89,379), 1.5% | NA | NA | 10015 (8474,11556) | 58 (-4,120), 0.6% | 87 (4,170), 0.9% | 222 (216,228) | 4 (-2,10), 1.8% | 12 (2,22), 5.4% |
| ≥ 85 | Before | 9772(9376,10168) | 12(5,19), 0.1% | 53(24,82), 0.5% | 172292 (165185,179399) | 582 (337,827), 0.3% | 14384 (13279,15489) | 242 (16, 468), 1.7% | 311 (14,608), 2.2% | 19486 (14383,24589) | 211 (169,253), 1.1% | 12512 (11305,13719) | 58 (53,63), 0.5% | 92 (81,103), 0.7% | 497 (371,623) | 1 (0,2), 0.2% | 33 (17,49), 6.6% |
|  | During | 8511 (±NA) | 26(±NA), 0.3% | 79(±NA), 0.9% | 178562 (166759,190365) | 1246 (52,2440), 0.7% | 12718 (10508,14928) | 184 (81, 287), 1.4% | 202 (71,333), 1.6% | NA | NA | 8616 (7341,9891) | 46 (-3,95), 0.5% | 66 (-10,142), 0.8% | 216 (193,239) | 4 (0,8), 1.9% | 10 (-1,21), 4.6% |
| **Additional age groups** |  |  |  |  |  |  |  |  |  |  |  |  |  |  |  |  |  |
| 18-49 | Before | NA | NA | NA | NA | NA | 5700 (5390,6010) | 45 (13,77) | 68 (15,121) | NA | NA | 8454 (8421,8487) | 30 (24,36), 0.4% | 51 (34,68), 0.6% | NA | NA | NA |
|  | During | NA | NA | NA | NA | NA | 5268 (4383,6153) | 57 (33,81) | 77 (43,111) | NA | NA | 5298 (3271,7325) | 18 (5,31), 0.3% | 31 (6,56), 0.6% | NA | NA | NA |
| 50-64 | Before | NA | NA | NA | NA | NA | 7455 (7099,7811) | 108 (30,186) | 141 (38,244) | NA | NA | 9146 (8932,9360) | 47 (44,50), 0.5% | 88 (67,109), 1% | 299 (223,375) | 2 (0,4), 0.7% | 14 (8,20), 4.7% |
|  | During | NA | NA | NA | NA | NA | 6308 (6225,6391) | 98 (45,151) | 117 (48,186) | NA | NA | 6596 (4853,8339) | 35 (15,55), 0.5% | 51 (18,84), 0.8% | 188 (165,211) | 3 (0,6), 1.6% | 6 (3,9), 3.2% |
| **Diagnosis groups** |  |  |  |  |  |  |  |  |  |  |  |  |  |  |  |  |  |
| Bronchitis&Bronchiolitis | Before | 412(358,466) | 18(10,26), 4.4% | 17(11,23), 4.1% | 9058(8881,9235) | 92 (70,114), 1% | 3362 (3054,3670) | 329 (78,580), 9.8% | 343 (79,607), 10.2% | 3363 (2522,4204) | 192 (157,227), 5.7% | 283 (266,300) | 16 (13,19), 5.7% | 11 (4,18), 3.9% | 129 (102,156) | 2 (1,3), 1.6% | 18 (7,29), 14% |
|  | During | 292(±NA) | 52(±NA), 17.8% | 44(±NA), 15.1% | 6204(5682,6726) | 168 (96,240), 2.7% | 1021 (922,1120) | 256 (148,364), 25.1% | 246 (132,360), 24.1% | NA | NA | 176 (139,213) | 9 (4,14), 5.1% | 8 (4,12), 4.5% | 34 (25,43) | 3 (3,3), 8.8% | 6 (3,9), 17.6% |
| Unspecified LRTI | Before | 517(341,693) | 2(±NA), 0.4% | 7(4,10), 1.4% | 175238(173822,176654) | 599 (379,819), 0.3% | 1117 (1046,1188) | 1 (-1,3), 0.1% | 6 (3,9), 0.5% | 1901 (1317,2485) | 67 (43,91), 3.5% | 20585 (19611,21559) | 86 (73,99), 0.4% | 164 (132,196), 0.8% | 159 (46,272) | NA | 20 (8,32), 12.6% |
|  | During | 297(±NA) | NA | 4(±NA), 1.3% | 122066(114133,129999) | 1025 (383,1667), 0.8% | 524 (511,537) | 0 (0,0), 0% | 5 (4,6), 1% | NA | NA | 11752 (10245,13259) | 64 (20,108), 0.5% | 106 (35,177), 0.9% | 44 (39,49) | 1 (±0), 2.3% | 2 (1,3), 4.5% |
| URTI | Before | 1616 (1499,1733) | NA | 5 (4,6), 0.3% | 49344 (48288,50400) | 127 (78,176), 0.3% | 3542 (3392,3692) | 3 (0,6), 0.1% | 32 (9,55), 0.9% | 12519 (9538,15500) | 208 (172,244), 1.7% | 3812 (3762,3862) | 22 (18,26), 0.6% | 32 (15,49), 0.8% | 30 (8,52) | NA | 7 (2,12), 23.3% |
|  | During | 1450 (±NA) | NA | 13(±NA), 0.9% | 38688 (33177,44199) | 155 (82,228), 0.4% | 1464 (1311,1617) | 2 (0,4), 0.1% | 18 (7,29), 1.2% | NA | NA | 2204 (1780,2628) | 10 (8,12), 0.5% | 10 (5,15), 0.5% | 2 (1,3) | NA | 1 (±0), 50% |
| Pneumonia&Influenza | Before | 41596 (39683,43509) | 49 (27,71), 0.1% | 220 (153,287), 0.5% | 440098 (435069,445127) | 1212 (837,1587), 0.3% | 45443 (43169,47717) | 354 (80,628), 0.8% | 556 (113,999), 1.2% | 87746 (69781,105711) | 652 (600,704), 0.7% | 30099 (28443,31755) | 119 (117,121), 0.4% | 232 (229,235), 0.8% | 831 (628,1034) | 5 (5,5), 0.6% | 26 (18,34), 3.1% |
|  | During | 33835 (±NA) | 111 (±NA), 0.3% | 328(±NA), 1% | 334450 (300266,368634) | 2055 (613,3497), 0.6% | 24347 (23874,24820) | 356 (207,505), 1.5% | 445 (250,640), 1.8% | NA | NA | 23661 (19622,27700) | 112 (26,198), 0.5% | 169 (46,292), 0.7% | 514 (487,541) | 10 (5,15), 1.9% | 13 (7,19), 2.5% |
| SARSCOV-2 | During | NA | NA | NA | 165462 (143401,187523) | 60 (13,107), 0% | 16491 (13517,19465) | 3 (2,4), 0% | 18 (7,29), 0.1% | NA | NA | 13026 (8287,17765) | 8 (8,8), 0.1% | 18 (10,26), 0.1% | 144 (53,235) | 1 (±0), 0.7% | 1 (±0), 0.7% |
| > 1 Diagnosis | Before | NA | NA | NA | 8406 (7912,8900) | 105 (93,117), 1.2% | 1266 (1155,1377) | 111 (23,199), 8.8% | 119 (27,211), 9.4% | 943 (791,1095) | 67 (59,75), 7.1% | NA | NA | NA | 23 (11,  35) | NA | 3 (1,5), 15% |
|  | During | NA | NA | NA | 106118 (81582,130654) | 398 (96,700), 0.4% | 5960 (5730,6190) | 94 (53,135), 1.6% | 98 (53,143), 1.6% | NA | NA | NA | NA | NA | 85 (35,135) | 2 (2,2), 2.4% | 1 (1,1), 1.2% |

**Main diagnosis group among RSV hospitalisations**

We categorised all RSV-coded and RSV-confirmed admissions by diagnosis group based on the recorded ICD codes at discharge. With few exceptions, pneumonia was the most common diagnosis group across age groups and seasons in all countries (35-100%), followed by bronchitis and bronchiolitis or unspecified LRTI, depending on the country and season (Figure S9). The proportion of URTI diagnoses was generally higher in the Netherlands and England compared to other countries. In Denmark, URTI diagnoses were more common when hospital admissions were RSV-confirmed rather than RSV-coded, a trend also observed in Spain-Valencia and Finland (Figure S9).

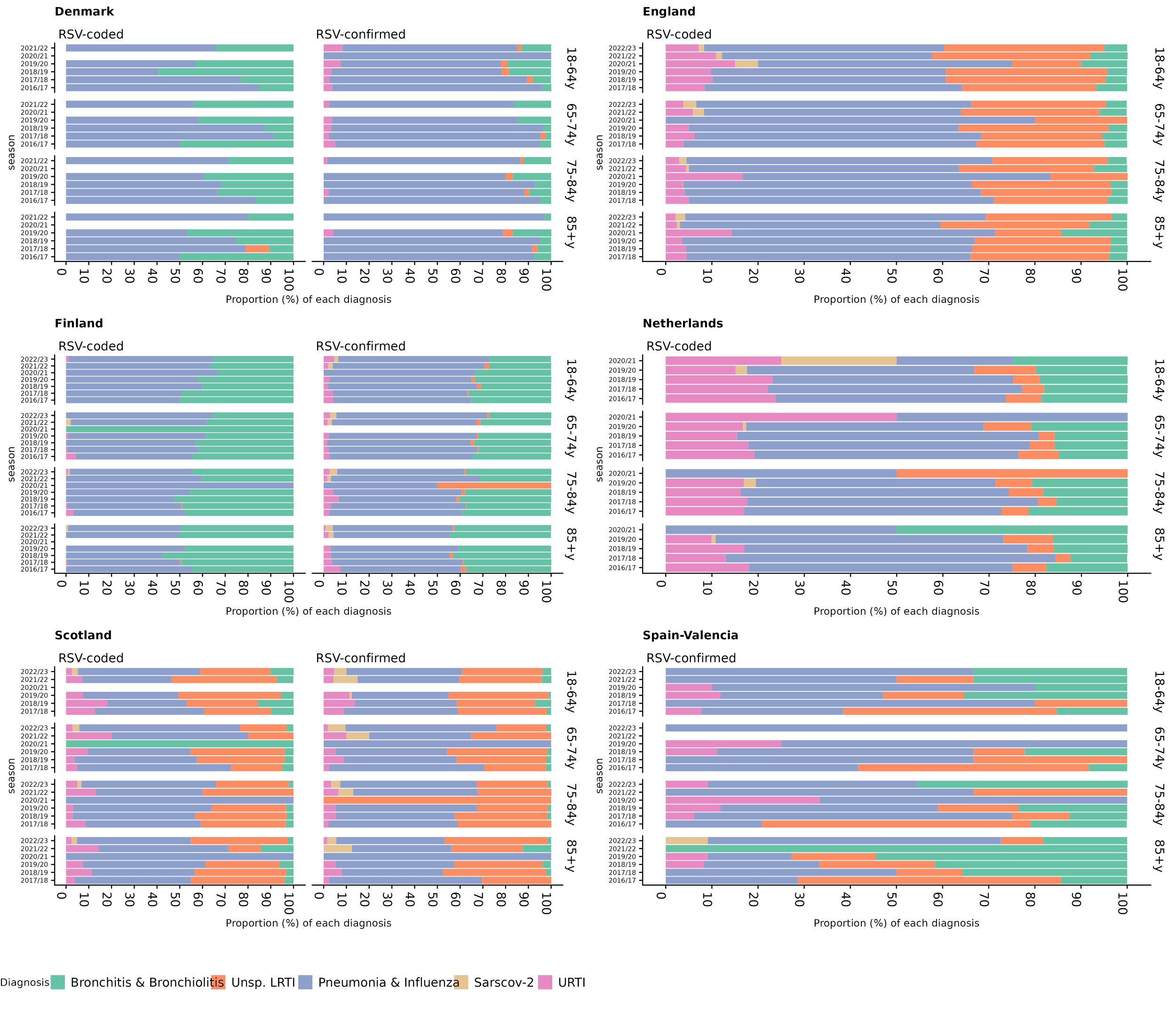

**Figure S1 Diagnosis groups among RSV-hospitalisations.** Each RSV-coded or RSV-confirmed hospitalisation was classified by the following diagnosis groups (Bronchitis & Bronchiolitis, Unspecified LRTI, Pneumonia & Influenza, SARSCOV-2 or URTI). The proportion in percentage (x-axis) of each of these diagnosis groups per country, season (y-axis), age group and hospitalisation type are shown.

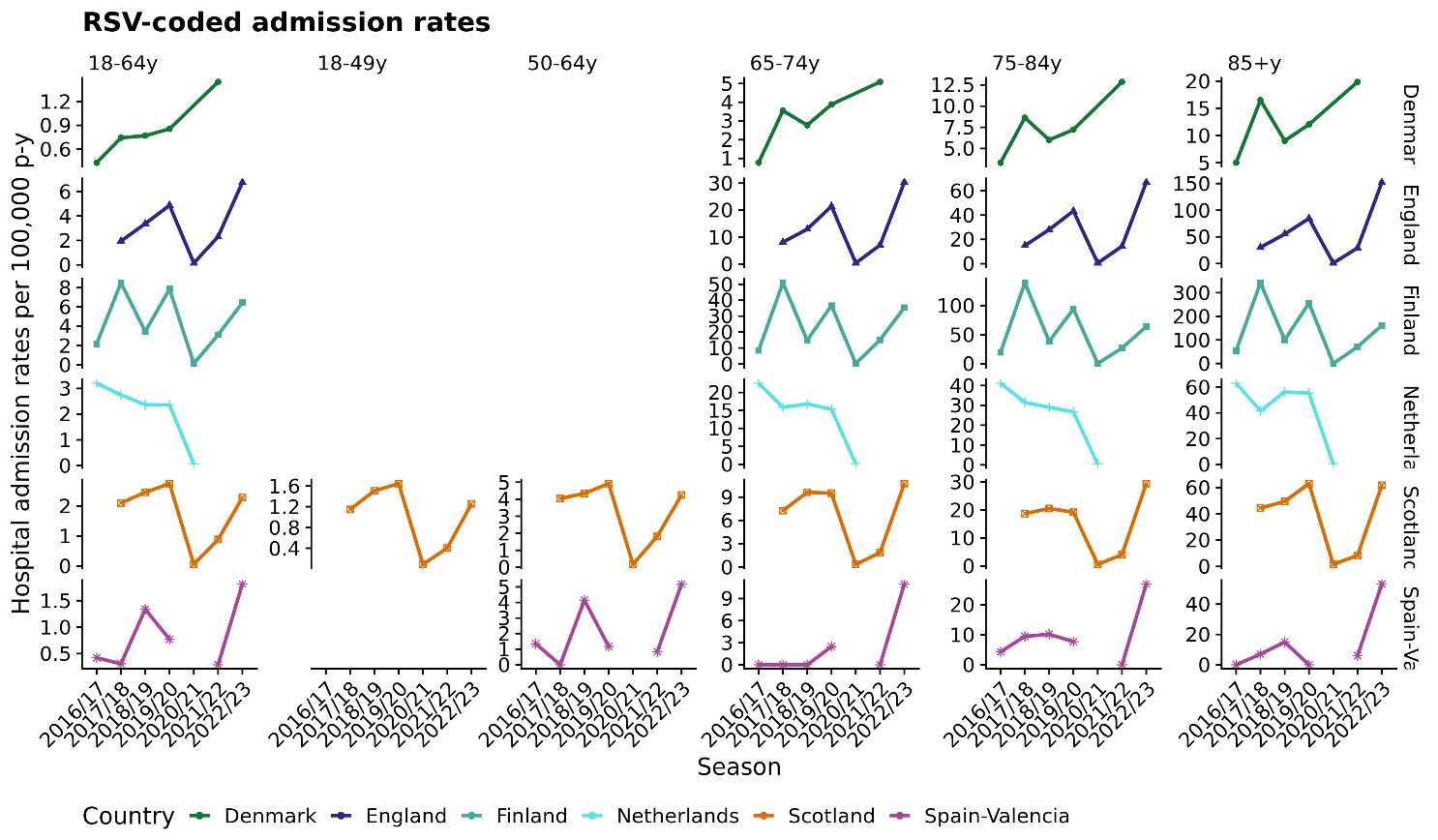
**Figure S2 RSV-coded hospitalisation admission rates (95%CI) in patients 18 years of age and above.** The RSV-coded hospital admissions rates per 100,000 persons-year (y-axis) and season (x-axis) are shown for each country (colour), age group (vertical subpanels) and hospitalisation type. Note that each panel has a different y-axis.

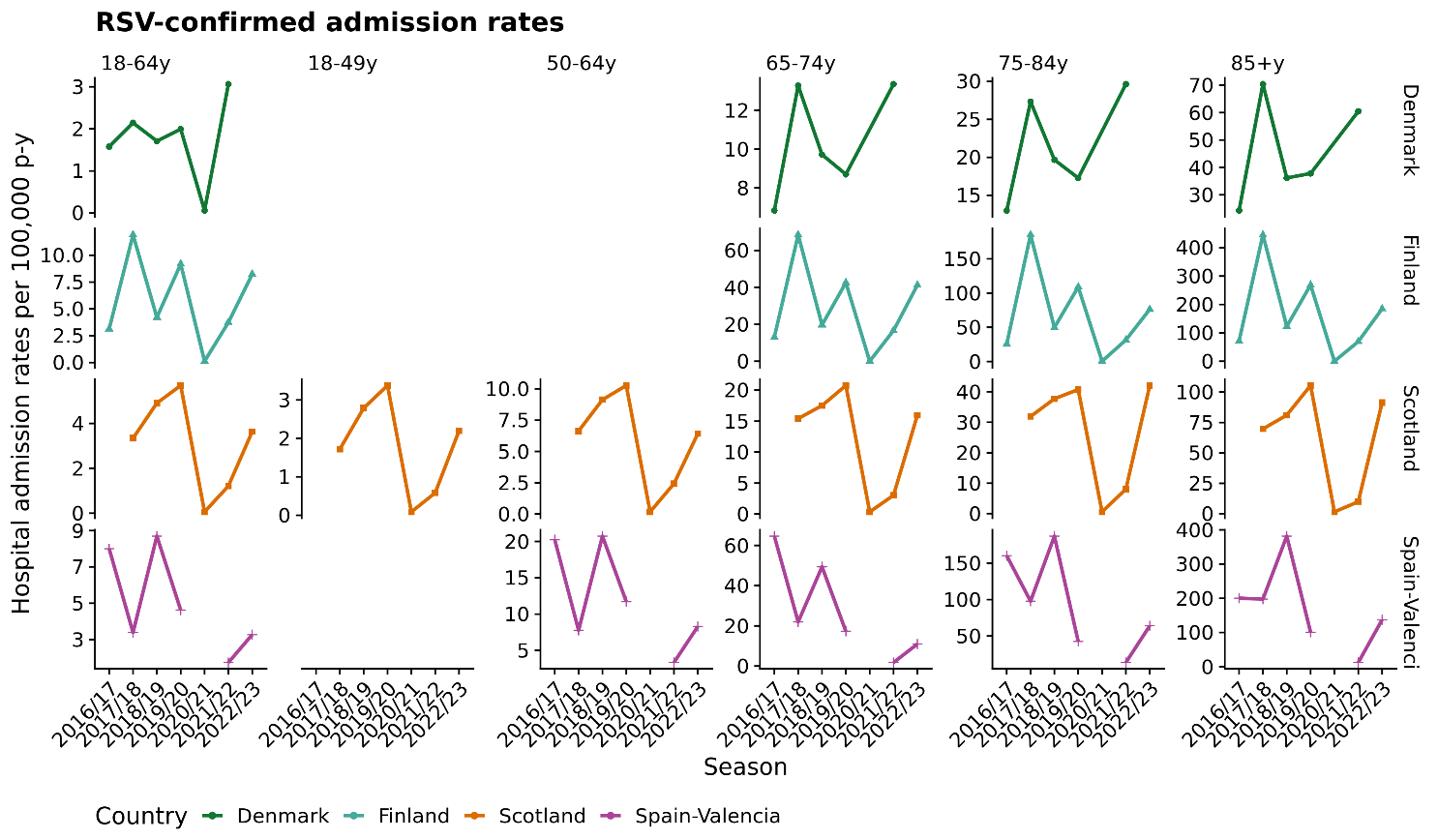

**Figure S3 RSV-confirmed hospitalisation admission rates (95%CI) in patients 18 years of age and above.** The RSV-confirmed hospital admissions rates per 100,000 persons-year (y-axis) and season (x-axis) are shown for each country (colour), age group (vertical subpanels) and hospitalisation type. Note that each panel has a different y-axis.

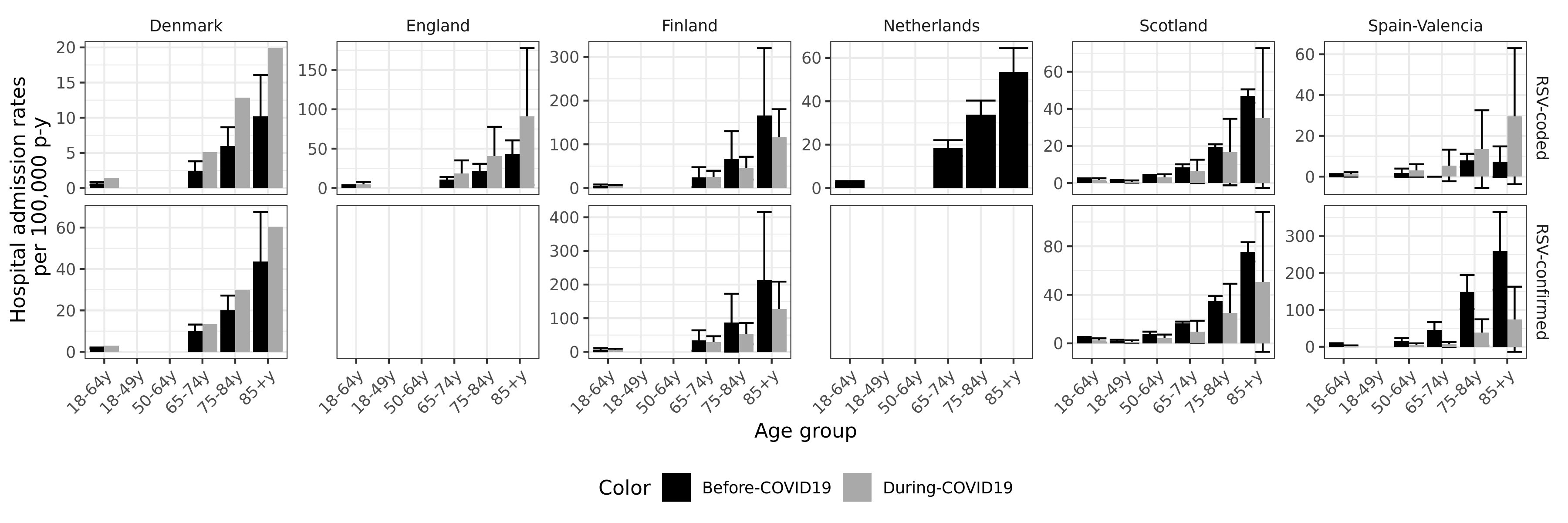
**Figure S4 Average hospital admission rates before and during COVID-19.** The average (±SD) of the hospital admission rates (y-axis) before COVID-19 seasons: 2016/17-2018/19 (black) and during COVID-19 seasons: 2021/22-2022(23 (grey) are shown for each country (horizontal subpanels, different colours) and age group (x-axis) for both RSV-coded and RSV-confirmed admissions.

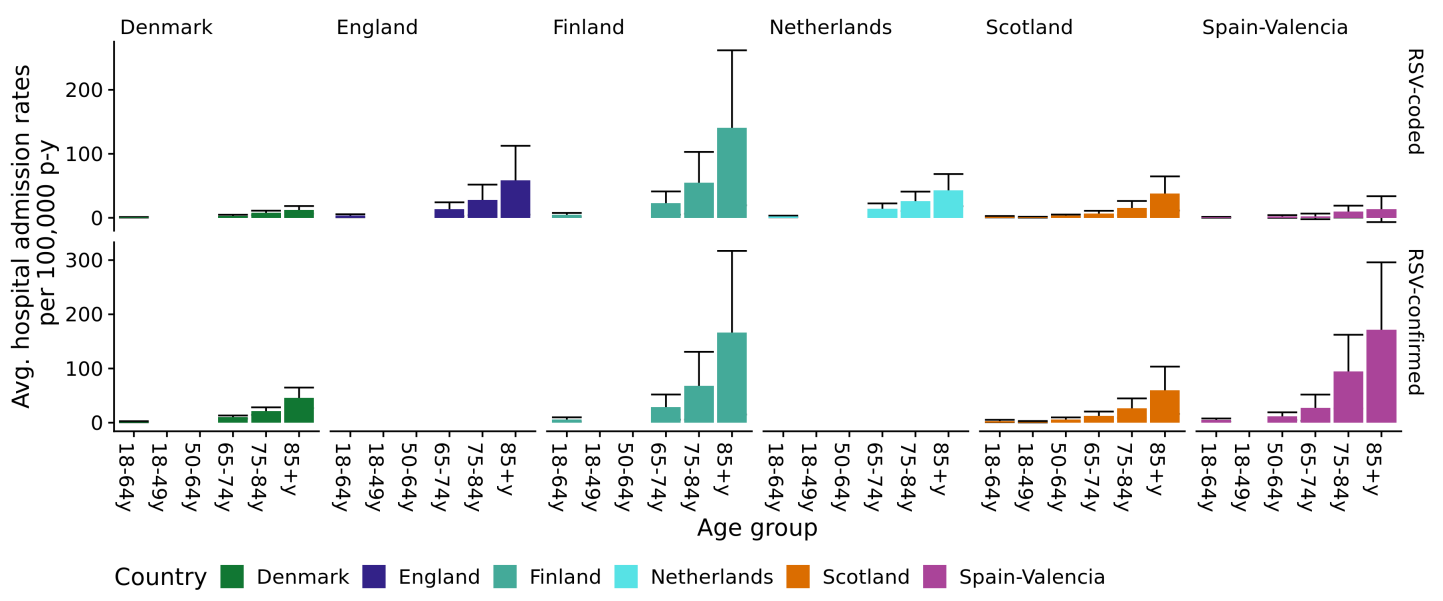

**Figure S5 Average hospital admission rates over the whole study period.** The average (±SD) of the hospital admission rates over the whole study period (y-axis) are shown for each country (horizontal subpanels, different colours) and age group (x-axis) for both RSV-coded and RSV-confirmed admissions.

**Table S3 RSV-coded admission rates (95%CI) per season, country and age group.** The RSV-coded admission rate for adults ≥18 years per 100,000 persons-year together with the 95%CI is shown per age group, country and season.

| **Surv. year** | **Age group** | **Denmark** | **England** | **Finland** | **Netherlands** | **Scotland** | **Spain-Valencia** |
| --- | --- | --- | --- | --- | --- | --- | --- |
| 2016/17 | 18-64y | 0.4 [0.2-0.7] | NA | 2.2 [1.7-2.7] | 3.2 [2.9-3.6] | NA | 0.4 [0-2.3] |
| 2017/18 |  | 0.7 [0.5-1.1] | 1.9 [1.8-2.1] | 8.5 [7.5-9.6] | 2.7 [2.4-3.1] | 2.1 [1.6-2.6] | 0.3 [0-1.7] |
| 2018/19 |  | 0.8 [0.5-1.1] | 3.4 [3.2-3.6] | 3.4 [2.8-4.1] | 2.4 [2.1-2.7] | 2.5 [2-3] | 1.3 [0.4-3.4] |
| 2019/20 |  | 0.9 [0.6-1.2] | 4.9 [4.6-5.1] | 7.9 [6.9-8.9] | 2.4 [2.1-2.7] | 2.7 [2.2-3.4] | 0.8 [0.1-2.8] |
| 2020/21 |  | NA | 0.1 [0.1-0.2] | 0.1 [0-0.3] | 0.1 [0-0.1] | 0.1 [0-0.2] | NA |
| 2021/22 |  | 1.4 [1.1-1.9] | 2.3 [2.1-2.5] | 3.1 [2.5-3.8] | NA | 0.9 [0.6-1.3] | 0.3 [0-1.6] |
| 2022/23 |  | NA | 6.7 [6.5-7] | 6.4 [5.6-7.4] |  | 2.3 [1.8-2.8] | 1.8 [0.6-4.2] |
| 2016/17 | 18-49y | NA | | | | NA | NA |
| 2017/18 |  |  |  |  |  | 1.1 [0.7-1.7] |  |
| 2018/19 |  |  |  |  |  | 1.5 [1-2.1] |  |
| 2019/20 |  |  |  |  |  | 1.6 [1.2-2.3] |  |
| 2020/21 |  |  |  |  |  | 0.1 [0-0.3] |  |
| 2021/22 |  |  |  |  |  | 0.4 [0.2-0.8] |  |
| 2022/23 |  |  |  |  |  | 1.3 [0.8-1.8] |  |
| 2016/17 | 50-64y | NA | | | | NA | 1.3 [0-7.5] |
| 2017/18 |  |  |  |  |  | 4 [2.9-5.4] | 0 [0-3.6] |
| 2018/19 |  |  |  |  |  | 4.3 [3.2-5.7] | 4.1 [1.1-10.6] |
| 2019/20 |  |  |  |  |  | 4.9 [3.7-6.4] | 1.2 [0-6.5] |
| 2020/21 |  |  |  |  |  | 0.2 [0-0.6] | NA |
| 2021/22 |  |  |  |  |  | 1.8 [1.1-2.8] | 0.8 [0-4.6] |
| 2022/23 |  |  |  |  |  | 4.3 [3.1-5.6] | 5.2 [1.7-12.1] |
| 2016/17 | 65-74y | 0.8 [0.3-1.8] | NA | 8.5 [6.4-11] | 22.6 [20.5-24.9] | NA | 0 [0-10.4] |
| 2017/18 |  | 3.6 [2.3-5.3] | 8.1 [7.4-8.9] | 50.9 [45.7-56.6] | 15.9 [14.1-17.8] | 7.3 [5.2-9.9] | 0 [0-7.4] |
| 2018/19 |  | 2.8 [1.6-4.4] | 12.9 [12-13.9] | 14.9 [12.1-18] | 16.8 [15-18.8] | 9.6 [7.3-12.5] | 0 [0-7.9] |
| 2019/20 |  | 3.9 [2.5-5.7] | 21.4 [20.3-22.7] | 36.6 [32.3-41.4] | 15.3 [13.6-17.2] | 9.5 [7.2-12.4] | 2.5 [0.1-13.8] |
| 2020/21 |  | NA | 0.4 [0.2-0.6] | 0.3 [0-1] | 0.1 [0-0.4] | 0.3 [0-1.2] | NA |
| 2021/22 |  | 5.1 [3.5-7.2] | 7 [6.3-7.8] | 15.1 [12.3-18.2] | NA | 1.8 [0.9-3.3] | 0 [0-6.5] |
| 2022/23 |  | NA | 30.3 [28.9-31.8] | 35.3 [31-40] | NA | 10.7 [8.3-13.7] | 11 [3.6-25.6] |
| 2016/17 | 75-84y | 3.3 [1.7-5.9] | NA | 19.6 [15.3-24.8] | 41.2 [37.2-45.4] | NA | 4.3 [0.1-24.1] |
| 2017/18 |  | 8.6 [5.8-12.3] | 14.9 [13.6-16.3] | 139.2 [127.2-152] | 31.5 [28.1-35.2] | 18.7 [14.3-24] | 9.5 [2-27.7] |
| 2018/19 |  | 6 [3.8-9.1] | 28 [26.2-29.9] | 38.4 [32.3-45.3] | 29 [25.8-32.5] | 20.5 [15.9-26] | 10.2 [2.1-29.8] |
| 2019/20 |  | 7.2 [4.8-10.4] | 43.3 [41.1-45.6] | 94.2 [84.6-104.6] | 26.7 [23.6-29.9] | 19.2 [14.8-24.5] | 7.7 [0.9-27.9] |
| 2020/21 |  | NA | 0.5 [0.3-0.8] | 0.5 [0.1-1.8] | 0.5 [0.1-1.1] | 0.6 [0.1-2.1] | NA |
| 2021/22 |  | 12.9 [9.7-16.7] | 14.3 [13.1-15.6] | 27 [22.3-32.5] | NA | 4 [2.2-6.8] | 0 [0-10] |
| 2022/23 |  | NA | 66.8 [64.1-69.6] | 63.8 [56.6-71.7] | NA | 29.4 [24-35.7] | 27 [11.6-53.1] |
| 2016/17 | ≥85y | 5 [1.8-10.9] | NA | 54.6 [43.2-68.1] | 62.9 [55.3-71.4] | NA | 0 [0-36.9] |
| 2017/18 |  | 16.6 [10.1-25.6] | 30.2 [27.3-33.3] | 342.1 [312.7-373.4] | 41.5 [35.4-48.3] | 44.3 [33.3-57.8] | 7.1 [0.2-39.3] |
| 2018/19 |  | 9 [4.5-16.2] | 55.3 [51.4-59.4] | 100.3 [84.8-117.8] | 56.2 [49.1-64] | 49.4 [37.8-63.5] | 15 [1.8-54.1] |
| 2019/20 |  | 12.1 [6.8-19.9] | 84.5 [79.7-89.4] | 255.6 [230.7-282.4] | 55.2 [48.3-62.9] | 62.9 [49.9-78.3] | 0 [0-31] |
| 2020/21 |  | NA | 0.9 [0.4-1.5] | 0 [0-2.4] | 0.5 [0.1-1.7] | 1.6 [0.2-5.6] | NA |
| 2021/22 |  | 19.9 [13-29.2] | 29.3 [26.5-32.3] | 70.4 [57.9-84.8] | NA | 8.4 [4.2-15] | 6.1 [0.2-33.8] |
| 2022/23 |  | NA | 152.3 [145.8-159] | 161.5 [142.4-182.5] | NA | 61.7 [49-76.7] | 53.3 [21.4-109.7] |

**Table S4 RSV-confirmed admission rates (95%CI) per season, country and age group.** The RSV-confirmed admission rate for adults ≥18 years per 100,000 persons-year together with the 95%CI is shown per age group, country and season.

| **Surv. year** | **Age group** | **Denmark** | **Finland** | **Scotland** | **Spain-Valencia** |
| --- | --- | --- | --- | --- | --- |
| 2016/17 | 18-64y | 1.6 [1.2-2.1] | 3.1 [2.6-3.8] | NA | 8 [4.8-12.5] |
| 2017/18 |  | 2.1 [1.7-2.7] | 11.9 [10.8-13.1] | 3.3 [2.8-4] | 3.4 [1.7-6.1] |
| 2018/19 |  | 1.7 [1.3-2.2] | 4.2 [3.5-5] | 4.9 [4.2-5.7] | 8.7 [5.7-12.7] |
| 2019/20 |  | 2 [1.6-2.5] | 9.2 [8.2-10.3] | 5.7 [4.9-6.6] | 4.6 [2.4-8.1] |
| 2020/21 |  | 0.1 [0-0.2] | 0.1 [0-0.3] | 0.1 [0-0.2] | NA |
| 2021/22 |  | 3.1 [2.5-3.7] | 3.7 [3.1-4.5] | 1.2 [0.9-1.6] | 1.7 [0.6-3.8] |
| 2022/23 |  | NA | 8.2 [7.3-9.3] | 3.6 [3-4.3] | 3.3 [1.5-6.2] |
| 2016/17 | 18-49y | NA | NA | NA | NA |
| 2017/18 |  |  |  | 1.7 [1.2-2.4] |  |
| 2018/19 |  |  |  | 2.8 [2.1-3.6] |  |
| 2019/20 |  |  |  | 3.4 [2.7-4.2] |  |
| 2020/21 |  |  |  | 0.1 [0-0.3] |  |
| 2021/22 |  |  |  | 0.6 [0.3-1] |  |
| 2022/23 |  |  |  | 2.2 [1.6-2.9] |  |
| 2016/17 | 50-64y | NA | NA | NA | 20.2 [11.3-33.4] |
| 2017/18 |  |  |  | 6.6 [5.2-8.3] | 7.8 [3.4-15.3] |
| 2018/19 |  |  |  | 9.1 [7.5-11.1] | 20.7 [12.7-32] |
| 2019/20 |  |  |  | 10.3 [8.5-12.3] | 11.7 [5.6-21.6] |
| 2020/21 |  |  |  | 0.2 [0-0.6] | NA |
| 2021/22 |  |  |  | 2.4 [1.6-3.5] | 3.3 [0.9-8.5] |
| 2022/23 |  |  |  | 6.4 [5-8.1] | 8.3 [3.6-16.3] |
| 2016/17 | 65-74y | 6.8 [5-9.2] | 13.1 [10.4-16.2] | NA | 64.6 [41-97] |
| 2017/18 |  | 13.3 [10.6-16.4] | 68.5 [62.4-75] | 15.4 [12.3-19] | 22 [11-39.3] |
| 2018/19 |  | 9.7 [7.5-12.4] | 19.6 [16.5-23.2] | 17.5 [14.2-21.3] | 49.4 [31.3-74.1] |
| 2019/20 |  | 8.7 [6.6-11.3] | 42.6 [37.9-47.7] | 20.8 [17.2-24.8] | 17.3 [7-35.7] |
| 2020/21 |  | NA | 0 [0-0.5] | 0.3 [0-1.2] | NA |
| 2021/22 |  | 13.3 [10.6-16.5] | 16.6 [13.8-19.9] | 3 [1.8-4.8] | 1.8 [0-9.8] |
| 2022/23 |  | NA | 41.2 [36.6-46.3] | 16 [12.9-19.5] | 11 [3.6-25.6] |
| 2016/17 | 75-84y | 13 [9.4-17.5] | 25.5 [20.5-31.3] | NA | 160.4 [112.9-221] |
| 2017/18 |  | 27.3 [22.1-33.4] | 185 [171.2-199.7] | 31.9 [26.1-38.6] | 97.8 [66.5-138.8] |
| 2018/19 |  | 19.7 [15.4-24.8] | 49.7 [42.7-57.5] | 37.7 [31.4-44.9] | 187.2 [141-243.7] |
| 2019/20 |  | 17.3 [13.4-21.9] | 108.9 [98.6-120.1] | 40.7 [34.2-48.1] | 42.5 [21.2-76] |
| 2020/21 |  | NA | 0.5 [0.1-1.8] | 0.6 [0.1-2.1] | NA |
| 2021/22 |  | 29.6 [24.7-35.2] | 31.5 [26.4-37.4] | 8.1 [5.4-11.7] | 13.6 [4.4-31.6] |
| 2022/23 |  | NA | 76.3 [68.4-84.9] | 42.1 [35.5-49.5] | 64 [38.5-100] |
| 2016/17 | ≥85y | 24.2 [16.2-34.8] | 71.4 [58.2-86.7] | NA | 200.3 [122.3-309.3] |
| 2017/18 |  | 70.4 [56.2-87] | 445.4 [411.8-481] | 69.8 [55.7-86.3] | 197.5 [131.2-285.4] |
| 2018/19 |  | 36.2 [26.3-48.6] | 122.6 [105.4-141.8] | 81.1 [65.9-98.6] | 381.7 [284.2-501.9] |
| 2019/20 |  | 37.8 [27.8-50.3] | 268.9 [243.3-296.3] | 105.4 [88.3-124.9] | 100.8 [52.1-176] |
| 2020/21 |  | NA | 0 [0-2.4] | 1.6 [0.2-5.6] | NA |
| 2021/22 |  | 60.5 [47.9-75.4] | 69.1 [56.8-83.4] | 9.9 [5.3-16.9] | 12.1 [1.5-43.8] |
| 2022/23 |  | NA | 184.8 [164.3-207.1] | 91.4 [75.8-109.3] | 136.9 [81.2-216.4] |

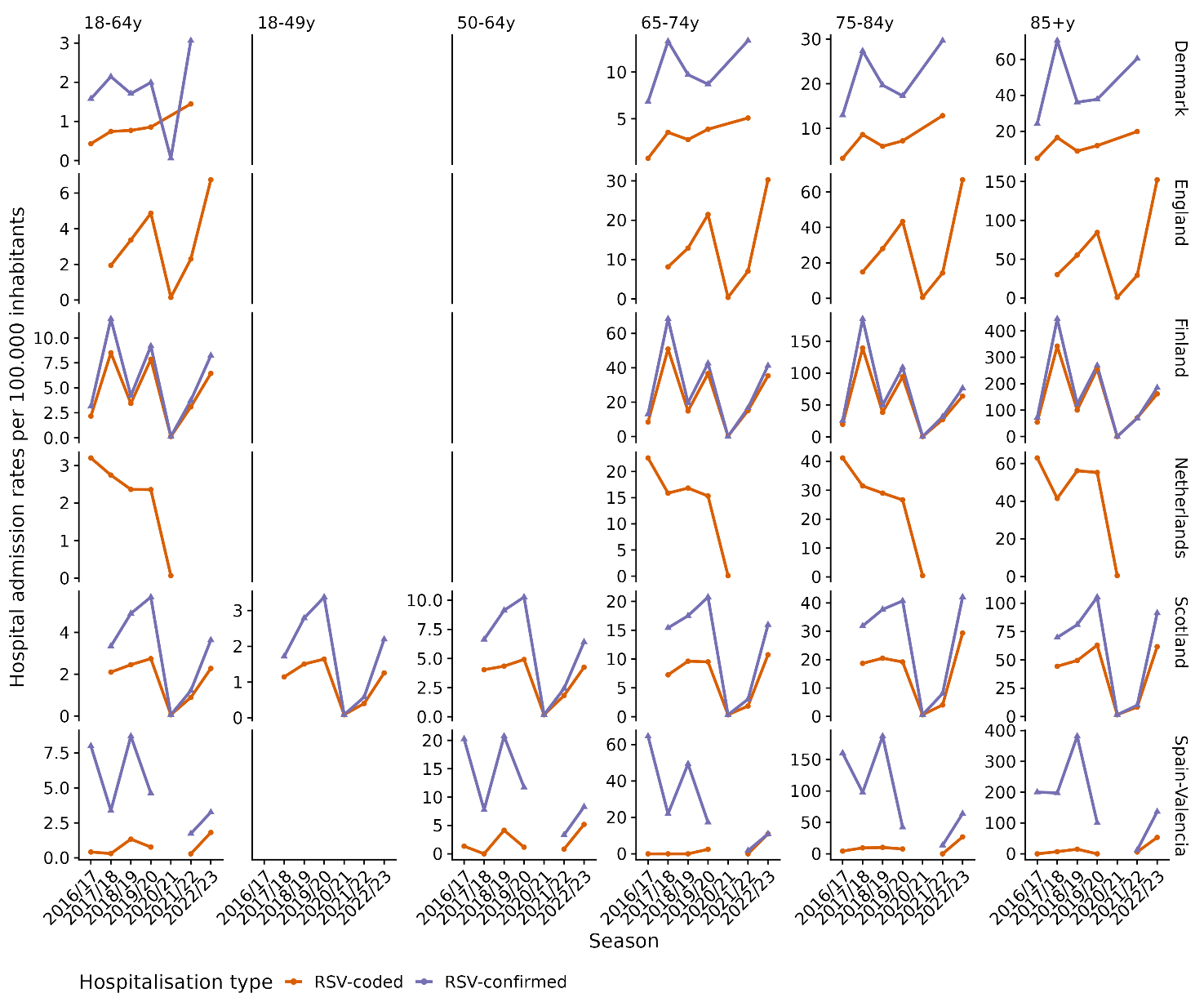
**Figure S6 RSV-coded and confirmed admissions per 100,000 person-years.** RSV-coded (orange) and RSV-confirmed (violet) admission rates are shown together for both RSV-coded and RS-confirmed admissions the same graph per country, age group and season.

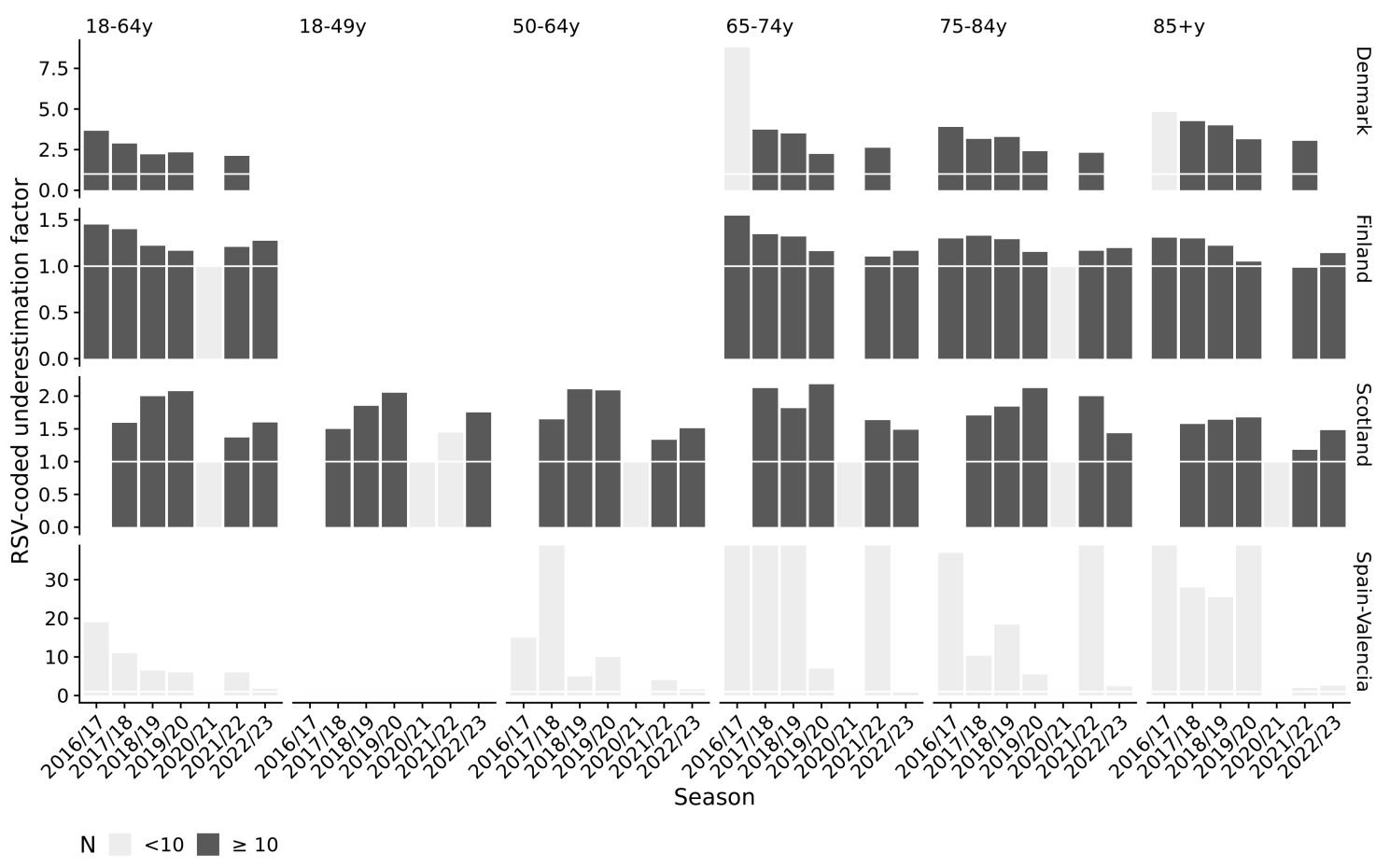

**Figure S7 RSV-coding practices and its underestimation.** The RSV-coded underestimation factor(y-axis) has been obtained by dividing in each season (x-axis), country (vertical subpanels) and age group (horizontal subpanels) the corresponding RSV-confirmed admission rate by the corresponding RSV-coded admission rate. Numbers above the white line indicate that the incidence estimated with RSV-coded admissions is lower than the one estimated with RSV-confirmed for the corresponding country, age group and season. Groups with less than 10 patients in the RSV-coded dataset are shown in light grey.

**Table S5 ICU admission rates.** Average (±SD) RTI and RSV-associated ICU admissions in England of the seasons before COVID-19 and of the seasons during COVID-19 stratified by age group and diagnosis group. The proportion of RTI/RSV-coded ICU admissions to the total number of RTI or RSV-coded admissions is also shown.

| **England** | | | |
| --- | --- | --- | --- |
| **Age group** | **COVID-19** | **RTI** | **RSV-coded** |
| 18-64 | Before | 12468 (12176,12760), 5.8% | 65 (42,88), 7.2% |
|  | During | 10318 (4931,15705), 3.8% | 47 (40,54), 3.0% |
| 65-74 | Before | 7372 (7040,7704), 6.1% | 46 (39,53), 7.9% |
|  | During | 4939 (2664,7214), 3.8% | 22 (19,25), 2.1% |
| 75-84 | Before | 6338 (6145,6531), 3.7% | 27 (16,38), 3.9% |
|  | During | 3716 (2336,5096), 1.9% | 12 (8,16), 0.9% |
| ≥ 85 | Before | 1996 (1899,2093), 1.2% | 5 (5,5), 0.9% |
|  | During | 882 (628,1136), 0.5% | 5 (4,6), 0.4% |
| **Diagnosis groups** |  |  |  |
| Bronchitis&Bronchiolitis | Before | 91 (89,93), 1% | 4 (3,5), 4.4% |
|  | During | 50 (44,56), 0.8% | 2 (1,3), 1.2% |
| Unspecified LRTI | Before | 3264 (3143,3385), 1.9% | 10 (9,11), 1.7% |
|  | During | 1280 (974,1586), 1.1% | 8 (7,9), 0.8% |
| URTI | Before | 420 (414,426), 0.9% | 2 (1,3), 1.6% |
|  | During | 201 (173,229), 0.5% | 1 (0,2), 0.7% |
| Pneumonia&Influenza | Before | 24053 (23948,24158), 5.5% | 97 (71,123), 8% |
|  | During | 11342 (9096,13588), 3.4% | 40 (32,48), 2.0% |
| SARSCOV-2 | During | 1914 (1354,2474), 1.2% | 1 (0,2), 1.7% |
| > 1 Diagnosis | Before | 321 (302,340), 3.8% | 6 (4,8), 5.7% |
|  | During | 5043 (1159,8927), 4.8% | 9 (9,9), 2.3% |

**Table S6 Case fatality rates.** Average (±SD) RTI and RSV-associated admissions that end up in death in England, Finland and Spain-Valencia of the seasons before COVID-19 and of the seasons during COVID-19 stratified by age group and diagnosis group. The case fatality rate to the total number of RTI or RSV-coded/confirmed admissions is also shown

|  |  | **England** | | **Finland** | | | **Spain-Valencia** | | |
| --- | --- | --- | --- | --- | --- | --- | --- | --- | --- |
| **Age group** | **COVID-19** | **Avg. RTI (±SD), CFR** | **Avg.**  **RSV-coded (±SD), CFR** | **Avg. RTI (±SD), CFR** | **Avg.**  **RSV-coded (±SD), CFR** | **Avg. RSV-confirmed (±SD), CFR** | **Avg. RTI (±SD), CFR** | **Avg.**  **RSV-coded (±SD), CFR** | **Avg. RSV-confirmed (±SD), CFR** |
| 18-64 | Before | 8391 (8159,8623), 3.9% | 30 (29,31), 3.3% | 481 (456,506), 3.7% | 4 (1,7), 2.8% | 5 (1,9), 2.5% | 6 (3,9), 1.2% | 0 (0,0), 0% | 0 (0,0), 0% |
|  | During | 10550 (9182,11918), 3.9% | 54 (12,96), 3.5% | 402 (175,629), 3.5% | 1 (0,2), 0.6% | 2 (2,2), 1% | 2 (2,2), 0.7% | 0 (0,0), 0% | 0 (0,0), 0% |
| 65-74 | Before | 12660 (12048,13272), 10.4% | 43 (40,46), 7.4% | 995 (967,1023), 8.6% | 7 (-1,15), 4% | 8 (-2,18), 3.5% | 13 (11,15), 3.3% | 0 (0,0), 0% | 1 (-1,3), 5.3% |
|  | During | 14282 (13243,15321), 10.9% | 86 (15,157), 8.2% | 958 (550,1366), 9.2% | 4 (2,6), 2% | 4 (0,8), 2.2% | 4 (2,6), 2.5% | 0 (0,0), 0% | 0 (0,0), 0% |
| 75-84 | Before | 24388 (22683,26093), 14.1% | 54 (40,68), 7.8% | 1910 (1767,2053), 12.2% | 13 (-1,27), 5.7% | 20 (-3,43), 6.5% | 27 (18,36), 5.0% | 0.3 (-1,1), 12.9% | 1 (0,2), 2.4% |
|  | During | 28196 (28170,28222), 14.6% | 152 (4,300), 10.8% | 1736 (1049,2423), 11.5% | 7 (0,14), 3.5% | 8 (0,16), 3.4% | 9 (5,13), 4.1% | 0 (0,0), 0% | 0 (0,0), 0% |
| ≥ 85 | Before | 34054 (30409,37699), 19.8% | 69 (49,89), 11.9% | 2559 (2396,2722), 17.8% | 30 (3,57), 12.3% | 38 (-4,80), 12.3% | 48 (26,70), 9.7% | 0.3 (-1,1), 30% | 4 (3,5), 11.12% |
|  | During | 35145 (34267,36023), 19.7% | 219 (-19,457), 17.6% | 2110 (1313,2907), 16.6% | 12 (3,21), 6.2% | 12 (1,23), 6.2% | 12 (8,16), 5.6% | 0.5 (-1,1), 12.5% | 2 (0,4), 15% |
| **Additional age groups** |  |  |  |  |  |  |  |  |  |
| 18-49 | Before |  |  | 69 (61,77), 1.2% | 1 (0,2), 2.9% | 1 (0,2), 1.9% |  |  |  |
|  | During |  |  | 72 (44,100), 1.4% | 0 (0,0), 0% | 1 (0,2), 1.3% |  |  |  |
| 50-64 | Before |  |  | 412 (381,443), 5.5% | 3 (1,5), 2.8% | 4 (1,7), 2.83% | 5 (3,7), 1.6% | 0 (0,0), 0% | 0 (0,0), 0% |
|  | During |  |  | 330 (131,529), 5.2% | 1 (0,2), 1.0% | 1 (0,2), 0.9% | 2 (2,2), 1.1% | 0 (0,0), 0% | 0 (0,0), 0% |
| **Diagnosis groups** |  |  |  |  |  |  |  |  |  |
| Bronchitis&Bronchiolitis | Before | 111 (100,122), 1.2% | 2 (1,3), 2.2% | 84 (75,93), 2.5% | 13 (3,23), 4% | 11 (2,20), 3.2% | 4 (3,5), 3.1% | 1 (NA,NA), 50% | NA |
|  | During | 113 (107,119), 1.8% | 6 (1,11), 3.6% | 26 (8,44), 2.5% | 4 (2,6), 1.6% | 4 (2,6), 1.6% | 2 (NA,NA), 5.9% | NA | NA |
| Unspecified LRTI | Before | 7393 (6868,7918), 4.2% | 16 (14,18), 2.7% | 51 (48,54), 4.6% | 0 (0,0), 0% | 0 (0,0), 0% | 5 (2,8), 3.1% | NA | 1 (NA, NA), 5% |
|  | During | 5505 (5371,5639), 4.5% | 35 (6,64), 3.4% | 27 (14,40), 5.2% | 0 (0,0), 0% | 0 (0,0), 0% | 1 (1,1), 2.3% | NA | NA |
| URTI | Before | 148 (143,153), 0.3% | 0 (-1,1), 0% | 44 (35,53), 1.2% | 0 (0,0), 0% | 1 (0,2), 3.1% | 2 (1,3), 6.7% | NA | NA |
|  | During | 124 (102,146), 0.3% | 4 (0,8), 2.6% | 18 (12,24), 1.2% | 0 (0,0), 0% | 1 (0,2), 5.6% | NA | NA | NA |
| Pneumonia&Influenza | Before | 71128 (67023,75233), 16.2% | 134 (112,156), 11.1% | 5651 (5369,5933), 12.4% | 32 (6,58), 9.0% | 50 (0,100), 9.0% | 39 (25,53), 4.7% | 1 (NA, NA) | 2 (1,3), 7.7% |
|  | During | 55010 (52307,57713), 16.5% | 304 (86,522), 14.8% | 2836 (1610,4062), 11.7% | 12 (5,19), 3.4% | 14 (5,23), 3.2% | 12 (6,18), 2.3% | NA | 1 (NA, NA), 7.7% |
| SARSCOV-2 | During | 8010 (7914,8106), 4.8% | 5 (0,10), 8.3% | 1459 (1300,1618), 8.9% | 0 (0,0), 0% | 4 (2,6), 22.2% | 12 (12,12), 8.3% | NA | NA |
| > 1 Diagnosis | Before | 680 (620,740), 8.1% | 12 (11,13), 11.4% | 113 (96,130), 8.9% | 8 (2,14), 7.2% | 8 (2,14), 6.7% | NA | NA | NA |
|  | During | 19330 (15179,23481), 18.2% | 75 (13,137), 18.8% | 842 (663,1021), 14.1% | 6 (2,10), 6.4% | 7 (4,10), 7.1% | 9 (9,9),  11% | NA | NA |

**Before and during COVID-19 statistical comparison**

*Statistical test*

To qualitatively analyse the change due to COVID-19, i.e., a difference observable since the start of the COVID-19 pandemic, we performed several statistical tests. First, we computed the average incidence rate by hospitalisation outcome (all hospitalisations, hospitalisation including an ICU stay, and hospitalisation leading to death), category (RTI-coded, RSV-coded, and RSV-confirmed), country and age group, for the before-COVID and since-COVID periods. The before-COVID period covered seasons from 2016 (2017 for England and Scotland) to 2019. The during-COVID period covered the 2021-2022 season for Denmark and Scotland and the 2021-2023 seasons for England, Finland, and Spain-Valencia. We considered 2019/2020 and 2020/2021 not to be representative, as strong NPI measures were in place and, therefore, were removed from this analysis. We computed a metric that we will call afterwards “ChangeProxy” to represent the change as the proportion of the difference between before- and during-COVID averages, compared to the before-COVID average period. We then leveraged Kruskal-Wallis test, a non-parametric test enabling comparisons of datasets whatever their distribution. We performed an initial series of nine tests to analyse if the ChangeProxy metric differed depending on the country, considering all age groups together but considering separately each hospitalisation outcome and category. ICU data being only present for England, the associated tests were withdrawn. Spain-Valencia data were not included in these tests as changes in hospital admission policies and procedures after COVID-19 may explain some of the differences observed. The effect of these changes has to be yet quantified before performing any statistical test. We then performed a second series of tests to directly assess a difference within the incidence rates between before- and during-COVID-19 periods, grouping the data by hospitalisation outcome, category, and age group.

The results of the analysis are available in Supplementary Figure 8. Kruskal-Wallis test considering all the age groups together and looking for differences between countries showed significant differences, for all the hospitalisation categories (RTI, RSV-coded or RSV confirmed) and severities (hospital admissions or deaths), except for RSV-confirmed deaths. When considering the incidences directly and taking all countries together, we observed no significant changes before and since COVID-19.

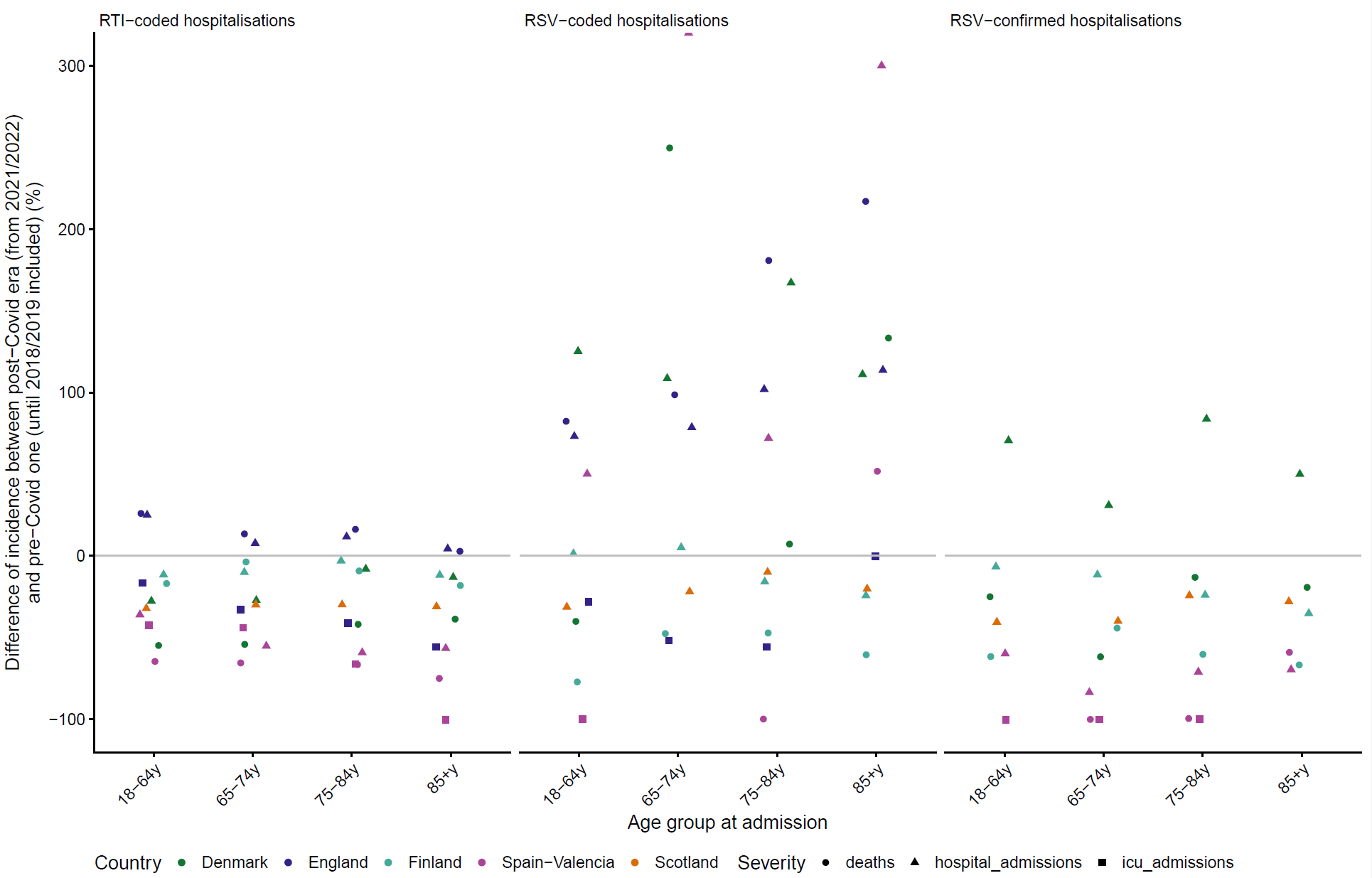

**Figure S8 Change of incidence of admissions before and during COVID-19 for patients 18 years or older,** stratified by category of hospitalisations (vertical subpanels, from left to right, RTI-coded, RSV-coded, and RSV-laboratory confirmed), coloured by country and shaped by severity of the hospital stay (hospitalisation only in general ward, including an ICU stay or leading to deaths).
