## Supplementary figures and images for "RSV healthcare burden in adults before and since the emergence of the COVID-19 pandemic in 6 European countries"

### Suppementary Fig 1

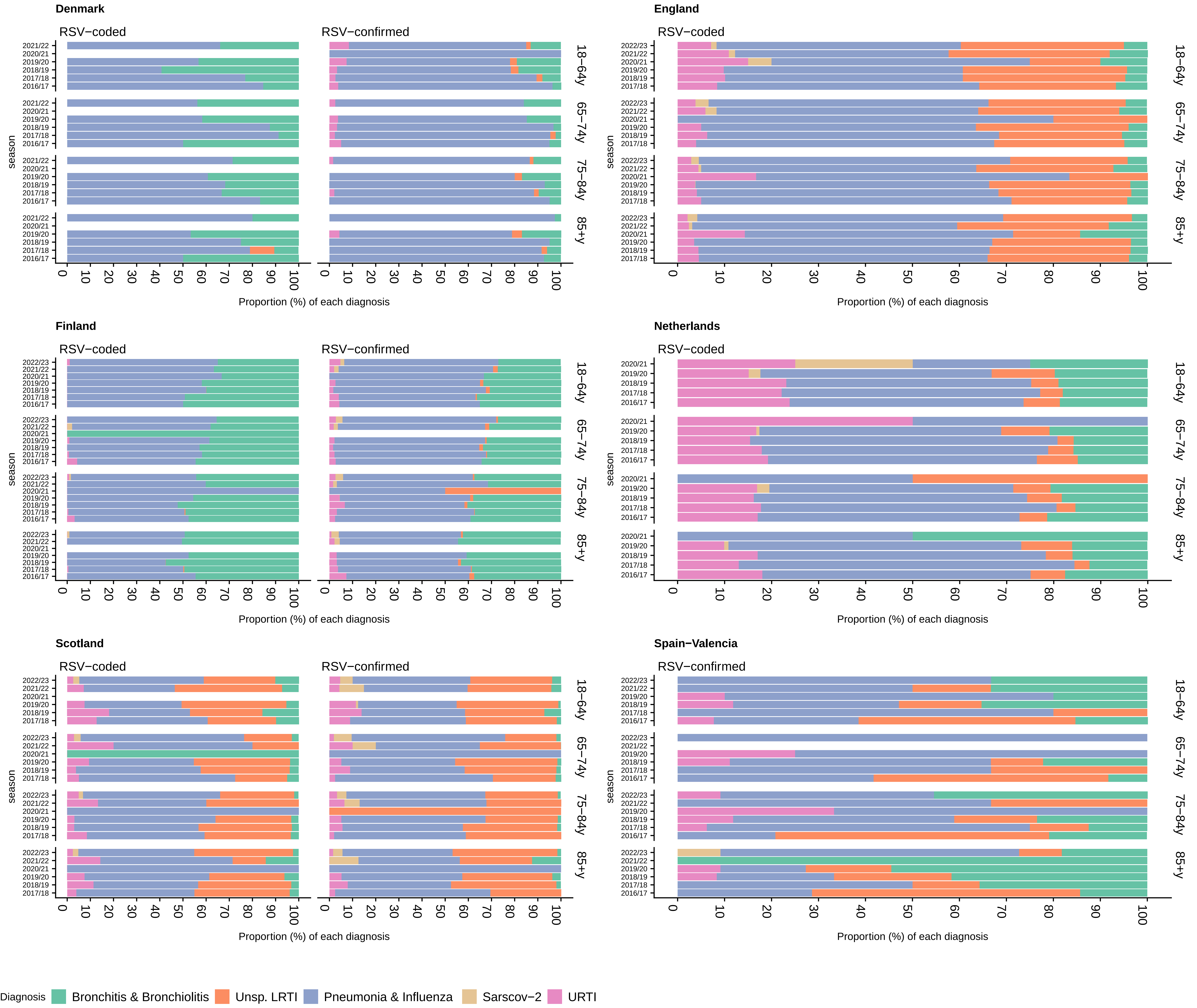

### Suppementary Fig 2

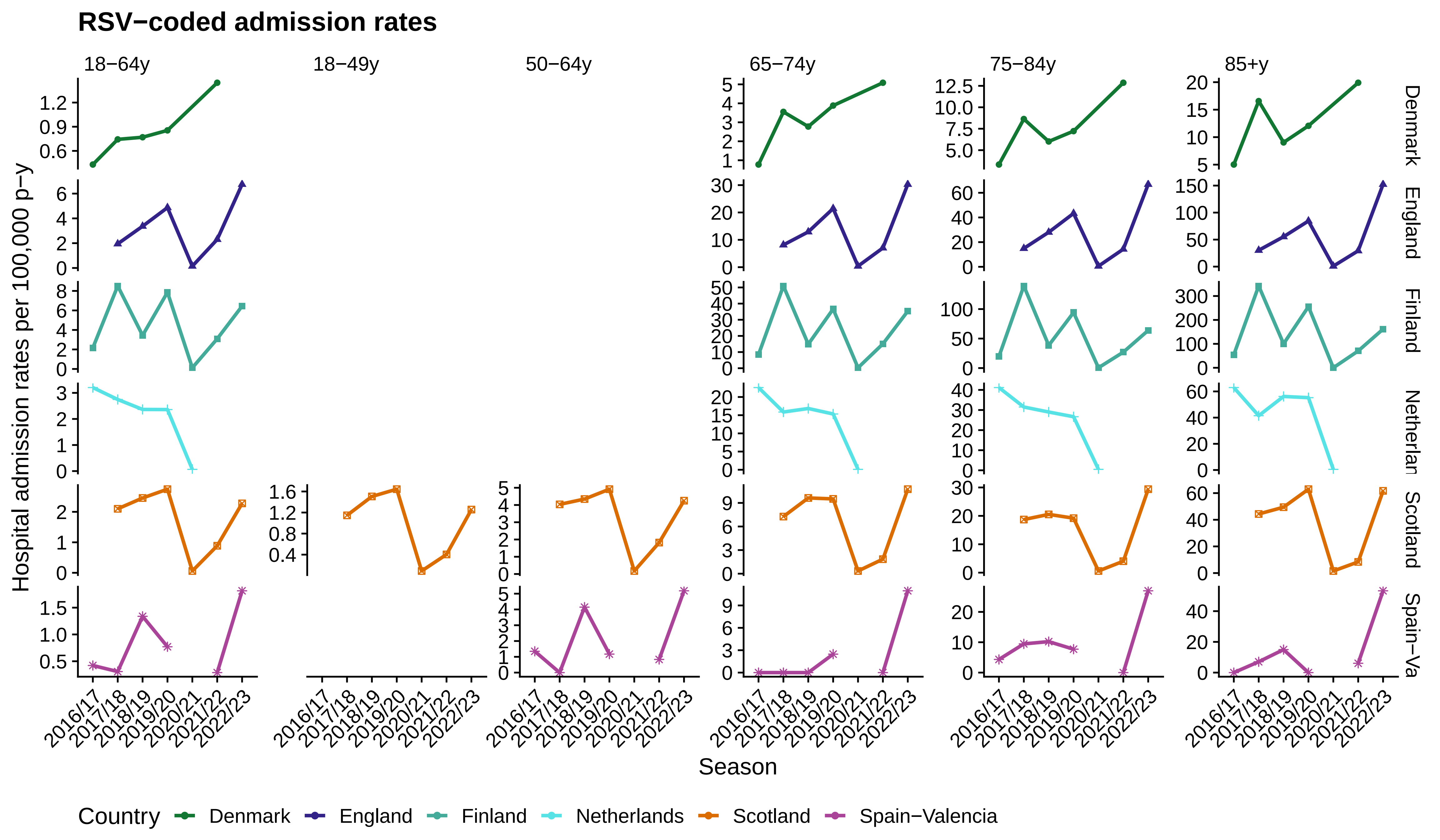

### Suppementary Fig 3

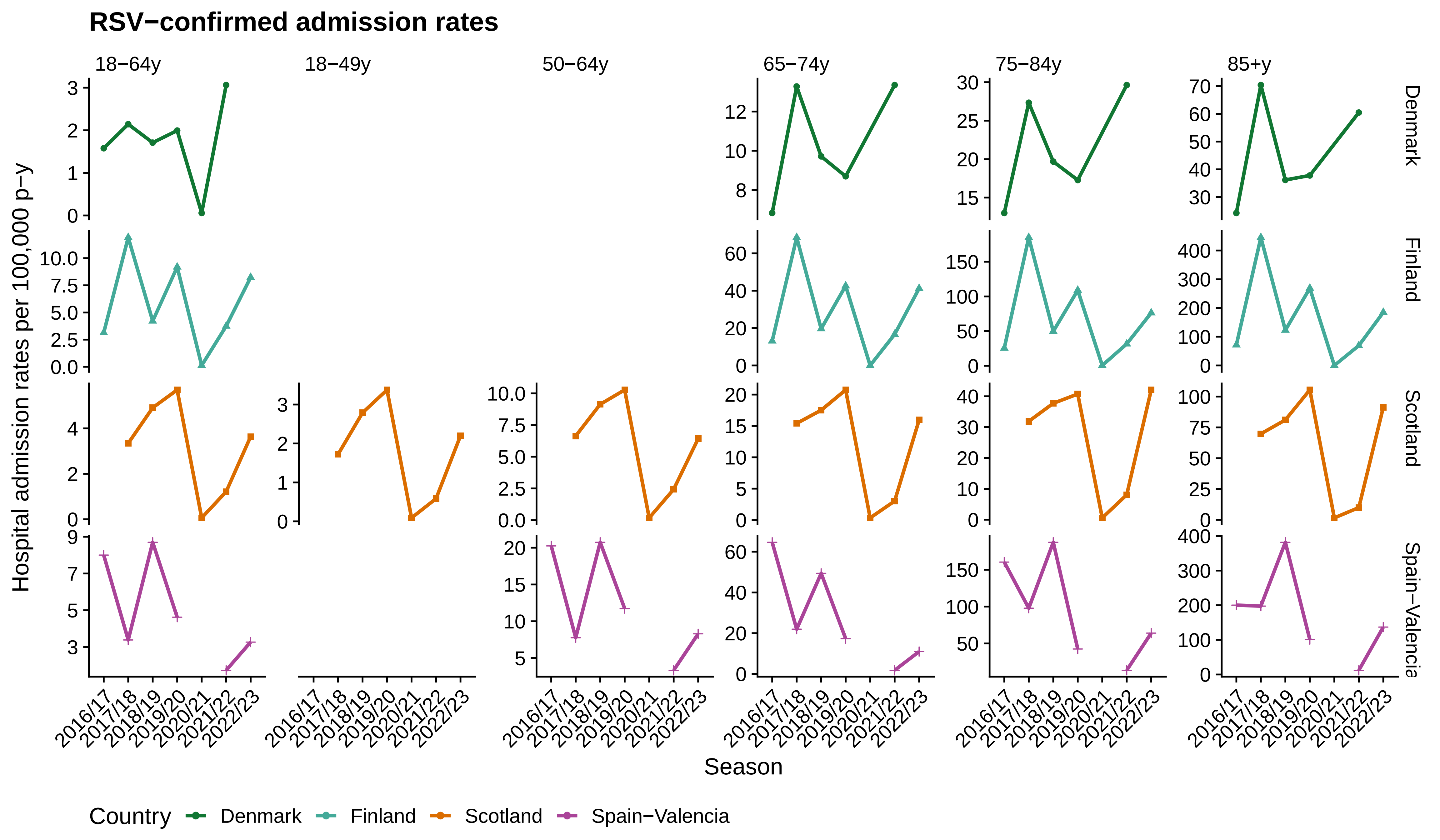

### Suppementary Fig 4

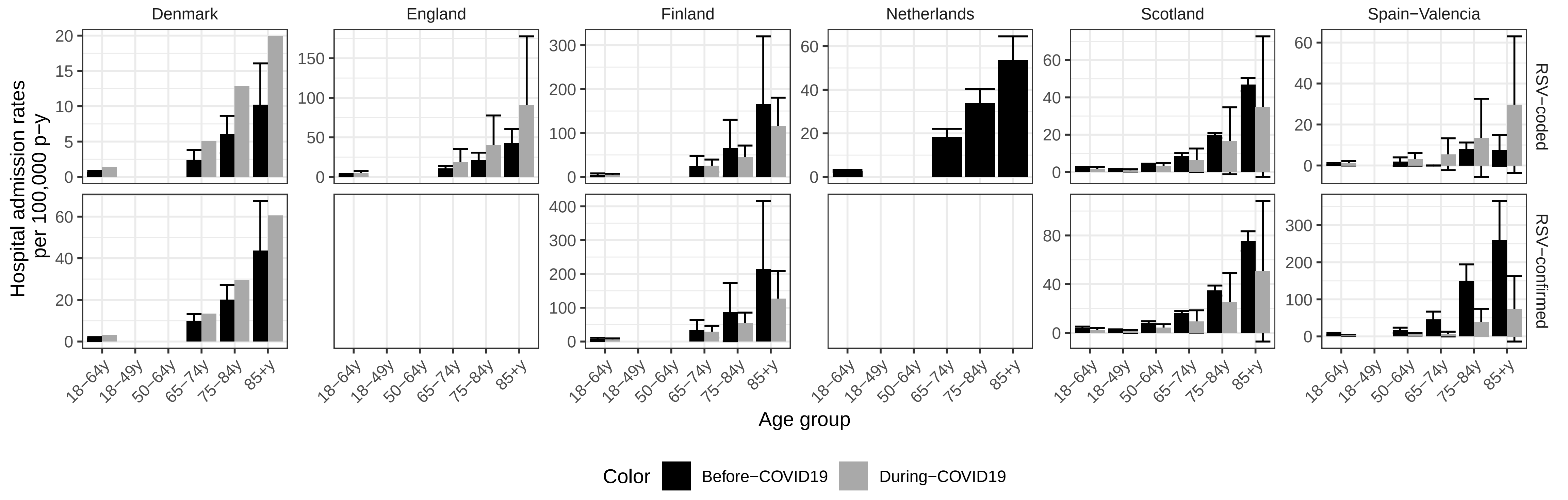

### Suppementary Fig 5

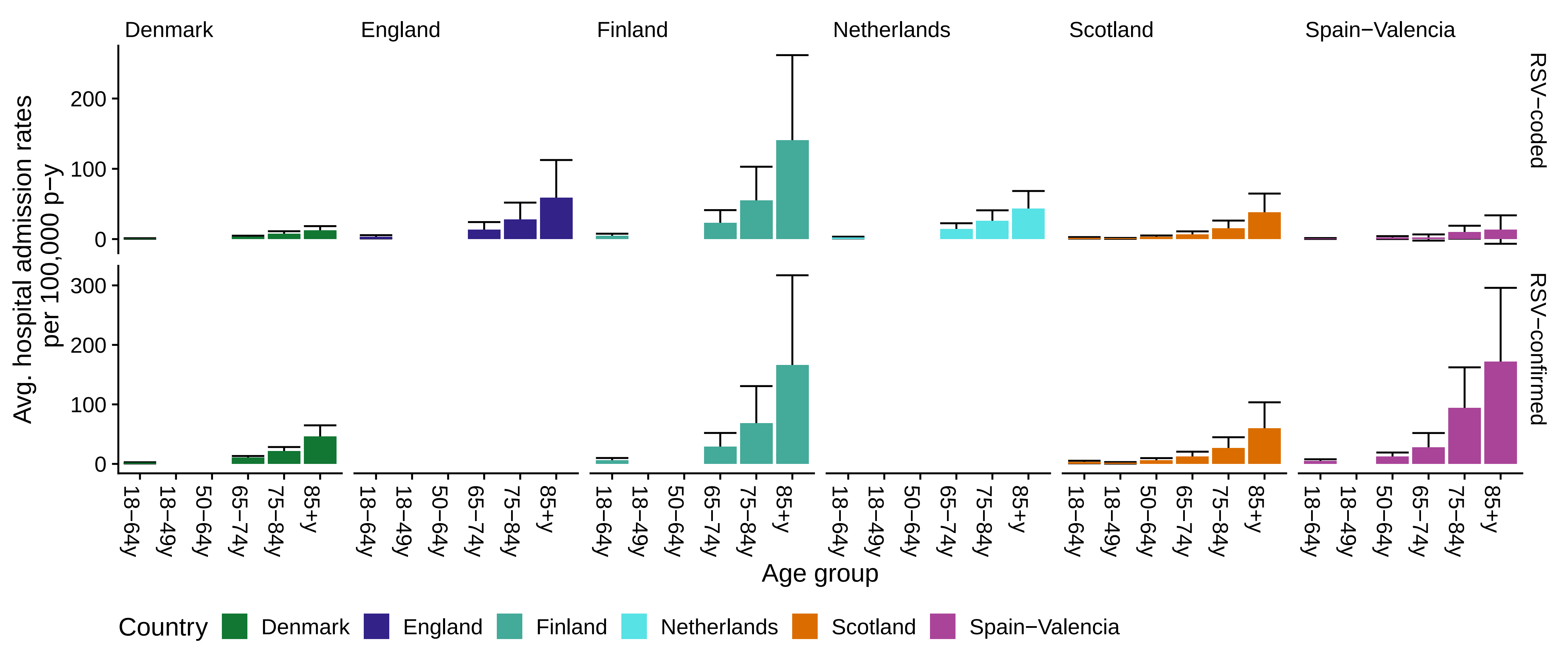

### Suppementary Fig 6

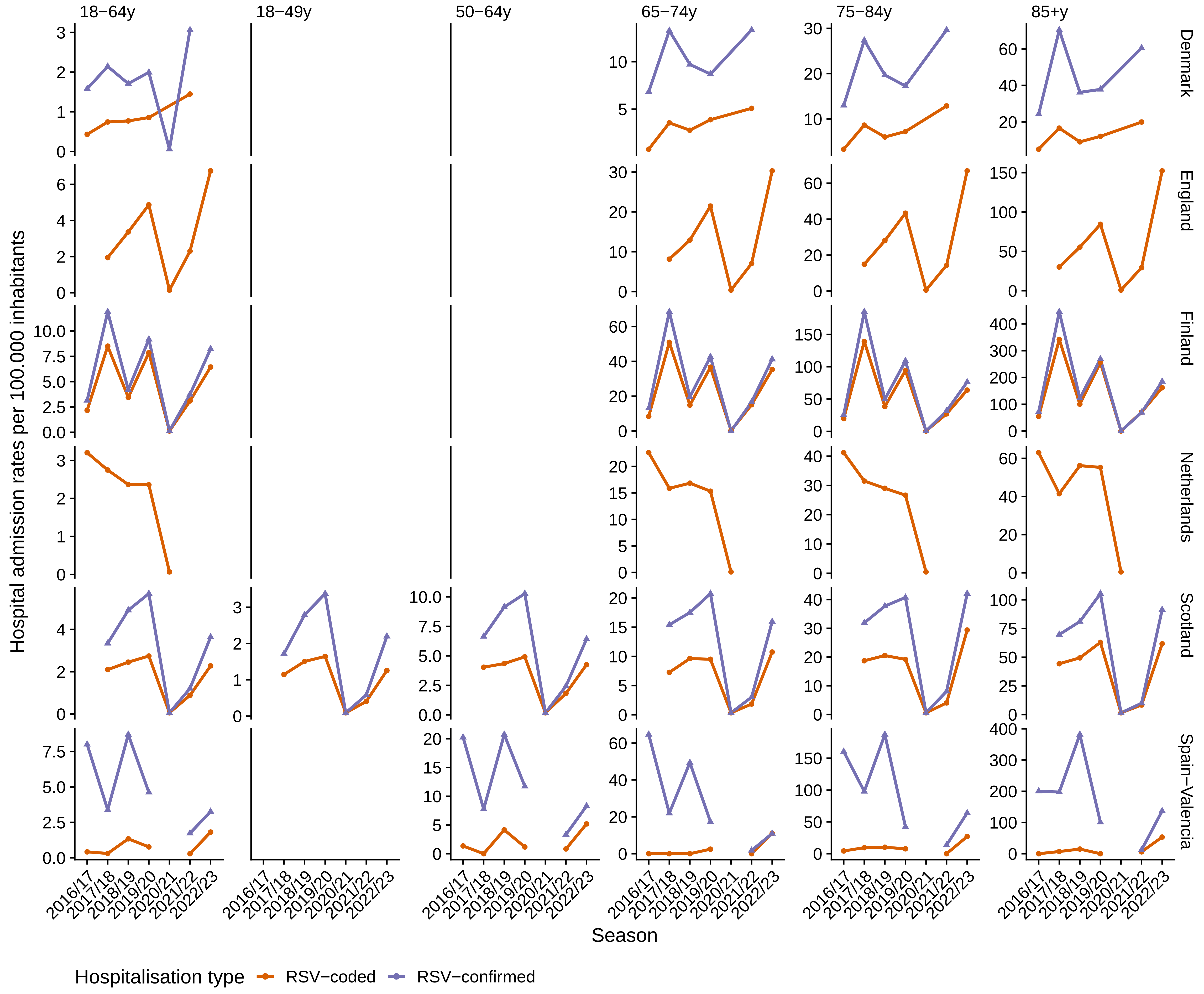

### Suppementary Fig 7

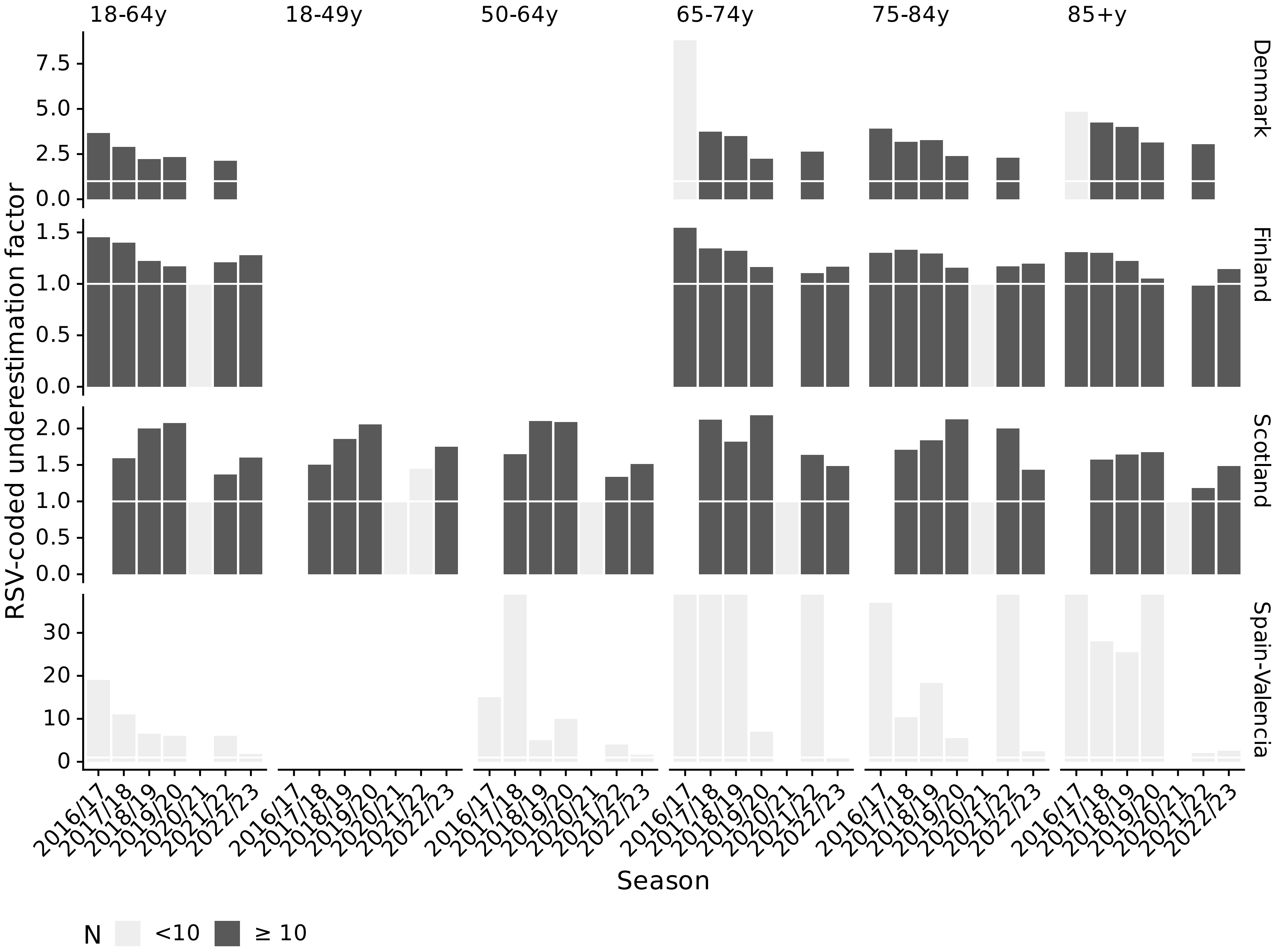

### Suppementary Fig 8

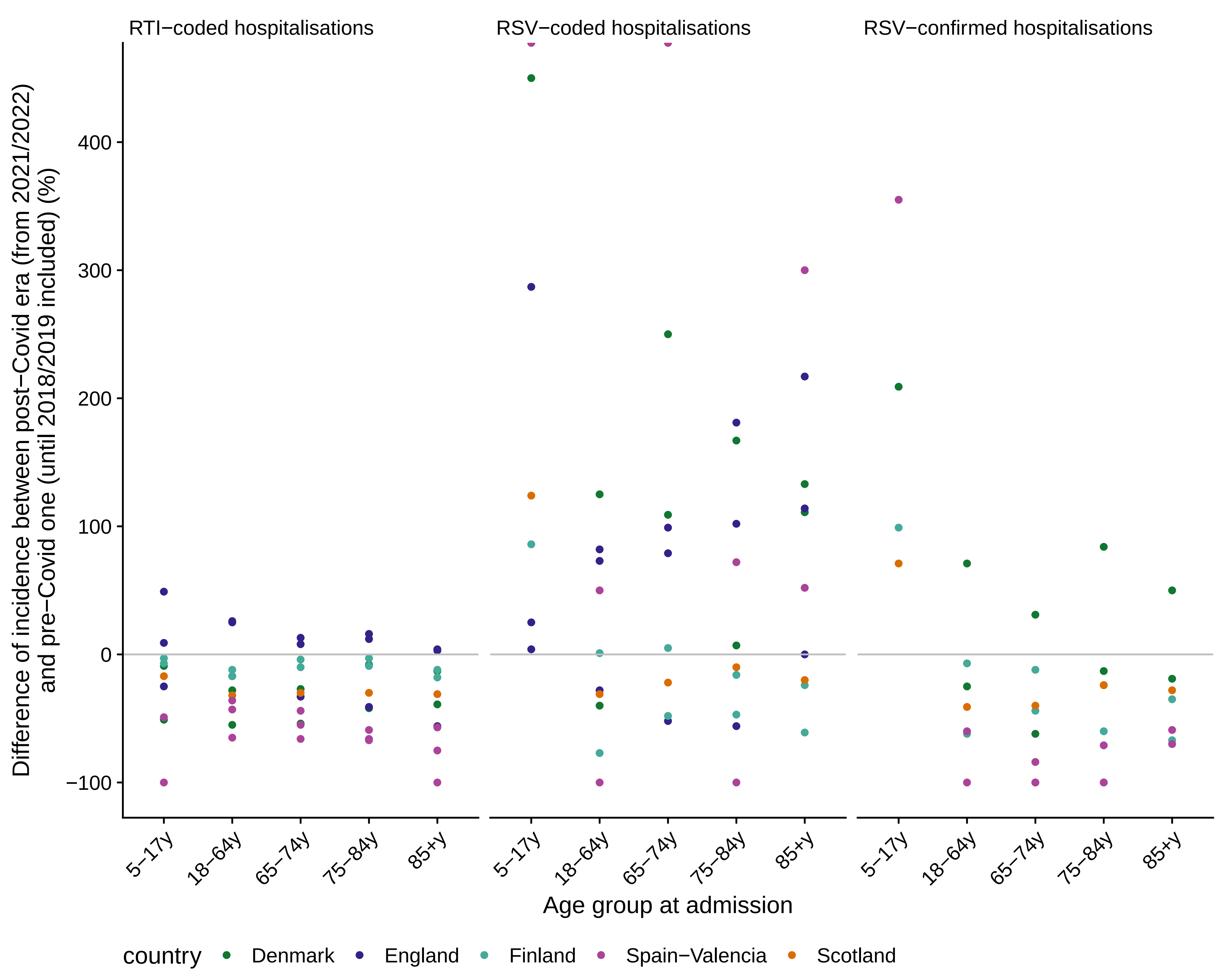
